# CD8^+^ T cell recall cytotoxicity during antiretroviral therapy is associated with limited HIV-1 reservoir size and activity

**DOI:** 10.1101/2025.10.10.25337688

**Authors:** David R. Collins, Mpho J. Olatotse, Jonathan M. Urbach, Zhu Zhuo, Suhui Zhao, Tyler J. Lilie, Rachel Raymond, Xianbao He, James Y. Chen, Bridget Coffey, Umar Arshad, Zachary J. Racenet, Hannah Wisner, Carlos Casquero, Xuan Guo, Lucy C. Walters, Jennifer M. Davis, Hannah C. Jordan, Noor Sohail, Kiera L. Clayton, Manish Sagar, James Billingsley, Shannan Ho Sui, Emanuele Mazzola, Bruce D. Walker, Athe Tsibris

**Affiliations:** Ragon Institute of Mass General Brigham, MIT and Harvard; Cambridge, MA, USA; Harvard Chan Bioinformatics Core, Harvard T.H. Chan School of Public Health, Boston, MA, USA; Division of Infectious Diseases, Brigham and Women’s Hospital; Boston, MA, USA; Department of Medicine, Boston Medical Center, Boston University Chobanian & Avedisian School of Medicine; Boston, MA, USA; Division of Infectious Diseases, University of Nebraska Medical Center; Omaha, NE, USA; Department of Pathology, University of Massachusetts Chan Medical School, Worcester, MA, USA; Department of Data Science, Dana-Farber Cancer Institute; Boston, MA, USA; Howard Hughes Medical Institute; Chevy Chase, MD, USA; Institute for Medical Engineering and Sciences and Department of Biology, Massachusetts Institute of Technology; Cambridge, MA, USA; Harvard Medical School; Boston, MA, USA

**Keywords:** HIV persistence, CD8^+^ T cells, analytical treatment interruption, human immunology

## Abstract

HIV-1 persists during antiretroviral therapy (ART), resulting in rebound viremia after treatment interruption. HIV-specific CD8^+^ T cell functionality is typically not restored by ART but may improve following prolonged treatment and was recently associated with post-intervention viral control. To evaluate the prevalence and impact of T cell immune function on HIV-1 persistence during prolonged ART, we mapped and functionally characterized CD8^+^ T cell responses that target virus reservoirs in sixty people with HIV-1 (PWH) who initiated therapy during chronic infection. Unexpectedly, 17% of participants exhibited robust proliferation and recall cytotoxicity against one or more autologous proviral epitopes at levels commensurate with spontaneous HIV-1 controllers, a group representing less than 1% of PWH. These functional responses were associated with smaller and less transcriptionally active HIV-1 reservoirs during ART. During an analytical treatment interruption, *in vivo* cytotoxic recall of phenotypically divergent HIV epitope-specific CD8^+^ T cell clonotypes trailed viral rebound but coincided with its attenuation prior to ART resumption. Our study reveals the unexpected prevalence of highly functional CD8^+^ T cells targeting autologous HIV-1 reservoirs among treated PWH and their potential impacts on viral persistence and rebound. These findings underscore a need for multimodal immunotherapeutic strategies capable of eliciting, restoring, or accelerating recall cytotoxicity toward achieving durable and scalable HIV-1 remission.

## INTRODUCTION

Cellular immune responses fail to control or eliminate infection in most people with HIV-1 (PWH) due to functional impairment of virus-specific CD8^+^ T cells and mutational escape from recognition^1^. In contrast, durable spontaneous control of HIV-1 infection is rare (∼1 in 300 PWH) and is associated with proliferative and cytolytic CD8^+^ T cell responses against non-escaped autologous viral epitopes^2, 3, 4, 5, 6^. Antiretroviral therapy (ART) is less commonly initiated during the earliest stages of HIV-1 infection but has been associated with preservation of HIV-specific CD8^+^ T cell proliferative capacity and memory formation^7, 8^, smaller persistent proviral reservoirs^9, 10, 11, 12, 13, 14, 15^, and a higher likelihood of post-treatment control of viremia^16, 17, 18, 19, 20^. In contrast, most PWH initiate ART during the chronic phase of HIV-1 infection; this typically fails to reverse functional impairment of virus-specific CD8^+^ T cells^21^. Recent evidence suggests that long-term ART can, in some instances, replenish HIV-specific CD8^+^ T cell function^22^ and that functional CD8^+^ T cells contribute to HIV-1 control following combination immunotherapy^23, 24, 25^. However, the prevalence of functional virus-specific CD8^+^ T cells in chronic treated HIV-1 infection and their impacts on HIV-1 persistence and rebound remain unclear.

Herein we examine virus-specific CD8^+^ T cell specificity, functionality, HIV-1 reservoir size, and treatment history in a cohort of 60 virologically suppressed PWH on prolonged ART initiated during chronic progressive HIV-1 infection. As expected, HIV-specific CD8^+^ T cells from most participants were either non-proliferative or failed to recognize autologous proviral epitopes due to mutational escape. However, HIV-specific CD8^+^ T cells in an unexpectedly large minority of individuals, over 50-fold more prevalent than spontaneous HIV-1 control, maintained robust proliferative and cytolytic potential against autologous proviral epitopes. Relative to participants with poorly functional and/or mutationally escaped CD8^+^ T cells, participants with highly functional, non-escaped responses harbored less total and intact HIV-1 DNA and significantly less cell-associated HIV-1 RNA. We characterized longitudinal immune responses in one participant who underwent ART interruption with close monitoring of viremia. Rebound viremia was accompanied by the sequential *in vivo* expansion of NK cells and cytolytic differentiation of phenotypically distinct HIV-specific CD8^+^ T cell clonotypes, which coincided with a rapid 3.9-fold viremic reduction prior to ART resumption. These findings demonstrate that HIV-specific CD8^+^ T cells with preserved recall cytotoxicity persist in a substantial fraction of PWH treated during chronic infection, comparable to the fraction of participants who achieved durable post-intervention control following broadly neutralizing antibody (bnAb) infusions in ART interruption trials^23, 25, 26, 27, 28, 29^. The association of recall cytotoxicity with smaller HIV-1 reservoirs and with attenuation of rebound viremia highlights the potential of inducing or enhancing such responses as part of multimodal immunotherapeutic strategies to elicit long-lasting ART-free HIV-1 remission.

## RESULTS

### CD8^+^ T cells can retain proliferative capacity against autologous HIV-1 during ART

HIV-1 epitope-specific CD8^+^ T cell responses were mapped using peripheral blood mononuclear cells (PBMCs) obtained from sixty PWH enrolled in the HIV Eradication and Latency (HEAL) study. All participants had initiated therapy during chronic progressive infection (median CD4 nadir 206 cells/mm^3^) and remained on ART for a minimum of one year (median 19 years) prior to sampling (Table 1). This study population was not enriched for protective or risk HLA genotypes known to have strong associations with viral load setpoint^30, 31^ (Extended Data Fig. 1a, Supplementary Data 1). HIV epitope-specific CD8^+^ T cell responses were mapped using interferon-γ (IFN-γ) enzyme-linked immunospot (elispot) following overnight stimulation with an array of individual HLA-optimal subtype B consensus HIV-1 peptides matched for each participant’s HLA genotype (Fig. 1a). 221 responses of varying magnitude were detected across 60 participants, with a median of 3 and a range of 0 to 18 epitope-specific responses per participant (Fig. 1b-c).

**Fig. 1:**
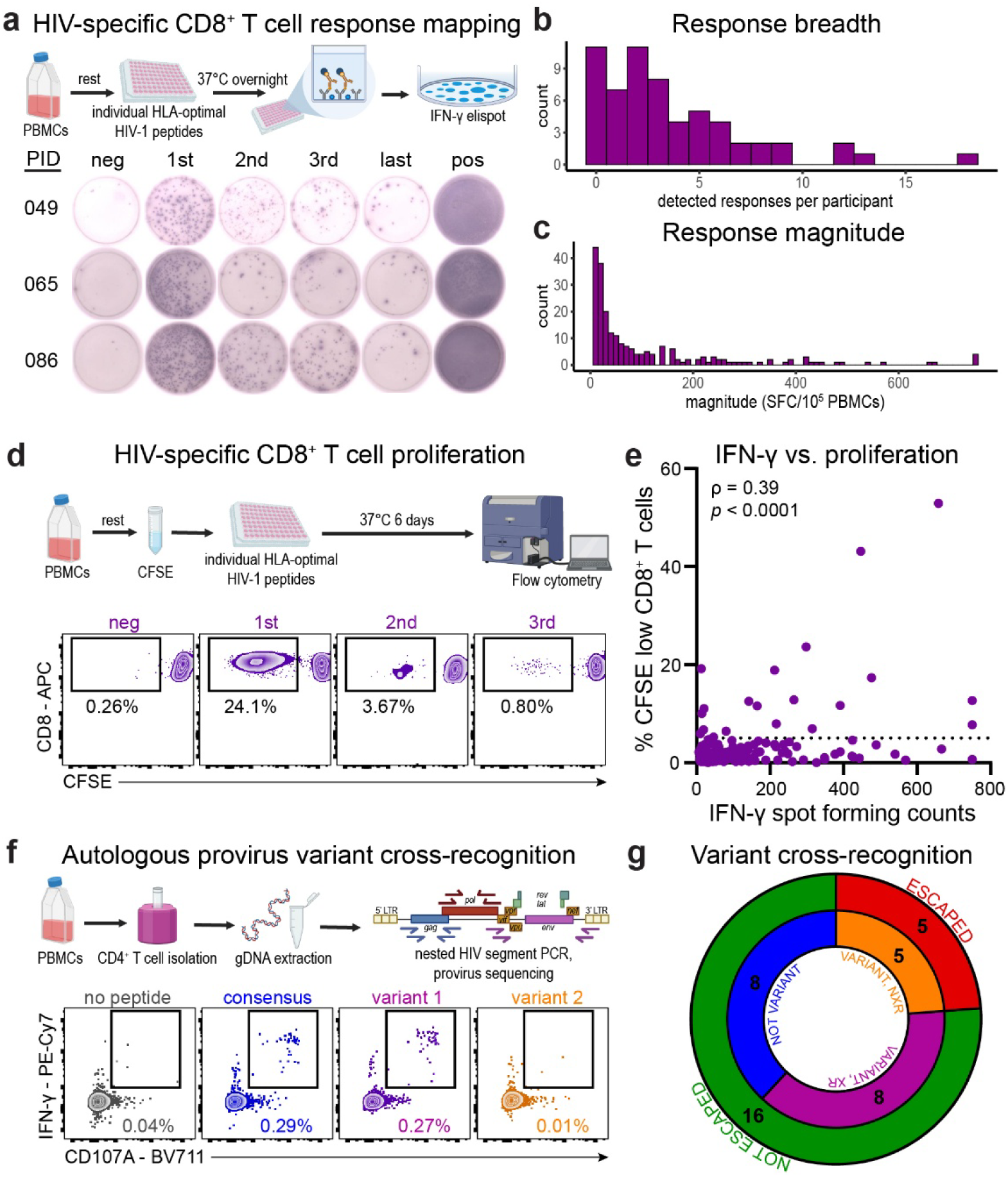
HIV-specific CD8^+^ T cell functionality against autologous proviral epitopes. **(a)** Schematic overview of HIV-specific CD8^+^ T cell response mapping screen and representative interferon-γ (IFN-γ) elispot results from 3 participants. **(b)** Histogram of CD8^+^ T cell response breadths measured by number of HLA-optimal HIV-1 epitopes eliciting IFN-γ elispot responses per participant (*n*=60). **(c)** Histogram of HIV-specific CD8^+^ T cell response magnitudes measured by IFN-γ spot-forming counts (SFC) per 100,000 PBMCs for each detectable response against HLA-optimal HIV-1 epitopes (*n*=221). **(d)** Schematic overview of HIV-specific CD8^+^ T cell proliferation assay and example results from the 3 largest responses detected by IFN-γ elispot in participant H003. **(e)** Summary of proliferation assay results across *n*=221 responses (from 60 participants) detected above background by IFN-γ elispot and Spearman correlation (ρ) with IFN-γ magnitudes as measured in c. Dashed line represents 5% proliferation, above which proliferation results were confirmed by independent replicates following initial screening. **(f)** Schematic overview of HIV-1 provirus sequencing and example cross-recognition assay results from participant H075 targeting consensus and two autologous proviral B*07 Nef TM9 variants. **(g)** Doughnut plot of autologous proviral epitope variant cross-recognition and mutational escape across *n*=21 proliferative responses (from 16 participants) identified in E. XR, cross-recognized; NXR, not cross-recognized.

**Table 1.** Demographics and clinical characteristics of study participants.

|  |  |  |
| --- | --- | --- |
| **Age at sampling** (years; median, IQR) |  | 54 (48–59) |
| **Sex** | Female (*n*, %) | 23 (38.3%) |
|  | Male (*n*, %) | 37 (61.7%) |
| **Race, Ethnicity** | Asian, Not Hispanic (*n*, %) | 1 (1.7%) |
|  | Black, Hispanic (*n*, %) | 2 (3.3%) |
|  | Black, Not Hispanic (*n*, %) | 19 (31.7%) |
|  | Black, Unknown (*n*, %) | 1 (1.7%) |
|  | White, Hispanic (*n*, %) | 7 (11.7%) |
|  | White, Not Hispanic (*n*, %) | 18 (30%) |
|  | White, Unknown (*n*, %) | 2 (3.3%) |
|  | Other/Unknown, Hispanic (*n*, %) | 10 (16.7%) |
| **HIV-1 diagnosis to ART initiation interval** (days; median, IQR) |  | 42 (4–479) |
| **ART initiation to sampling interval** (years; median, IQR) |  | 19 (11–22) |
| **Peak plasma viral load** (log<sub>10</sub> HIV-1 RNA copies/ml; median, IQR) |  | 4.9 (3.9–5.5) |
| **Viral load at sampling** (HIV-1 RNA copies/ml) |  | Undetectable (<20) |
| **CD4 nadir** (cells/mm<sup>3</sup>; median, IQR) |  | 202 (27–367) |
| **CD4 count at sampling** (cells/mm<sup>3</sup>; median, IQR) |  | 689 (511–913) |

We next assessed virus-specific CD8^+^ T cell functionality by measuring HIV-1 epitope-specific CD8^+^ T cell proliferation via CFSE dilution in response to six-day peptide stimulation (Fig. 1d). As expected, the proliferative capacity of most HIV-specific responses detected by elispot was severely impaired, with only a modest correlation between IFN-γ production and proliferation (Fig. 1e). Unexpectedly, however, 21 (9.5%) of 221 HIV-1 epitope-specific responses from 17 of 60 HEAL participants exhibited proliferative capacity surpassing the median observed among spontaneous controllers (≥5% CFSE dilution, Extended Data Fig. 1b-d), who represent less than 1% of PWH.

Viral mutational escape from T cell recognition is one potential mechanism by which CD8^+^ T cell functionality may be maintained^32^. To examine whether these highly proliferative responses recognized cognate epitope variants present within autologous HIV-1 proviruses, we sequenced targeted epitopes within proviral DNA using nested PCR of segmented amplicons from CD4^+^ T cells and then measured degranulation and cytokine production in response to stimulation with peptides matching each autologous epitope sequence observed (Fig. 1f). Analysis of proviral sequences revealed autologous variants relative to the clade B consensus within epitopes targeted by 13 of the 21 observed highly proliferative responses. Of these, 5 had escaped from recognition and 8 were efficiently cross-recognized, totaling 16 non-escaped, highly proliferative responses from 14 participants (Fig 1g). These results identified a subset of PWH with highly proliferative CD8^+^ T cell responses to autologous proviral epitopes following prolonged ART initiated during chronic progressive infection.

### Proliferative HIV-specific CD8^+^ T cells durably maintain high recall cytotoxicity

Using an expanded antigen-specific elimination assay^33^, we next measured the capacity of immunodominant HIV-1 epitope-specific CD8^+^ T cells to kill autologous peptide-pulsed CD4^+^ T cells after a six-day *in vitro* re-exposure to antigen (Fig. 2a). Proliferative responses exhibited higher recall cytotoxicity than nonproliferative responses (Fig. 2b) and we observed a strong positive correlation between proliferation and recall cytotoxicity (Fig. 2c). Of the 16 non-escaped highly proliferative responses we observed from 14 participants, 11 proliferative responses from 10 participants (17% of the cohort) demonstrated recall cytotoxicity greater than the first quartile of immunodominant CD8^+^ T cell responses from spontaneous controllers (Extended Data Fig. 1e). Responses in HEAL participants with high proliferation and recall cytotoxicity were not unique in their HLA restriction or HIV-1 protein/epitope targeting (Extended Data Fig. 1f-g, Supplementary Data 1).

**Fig. 2:**
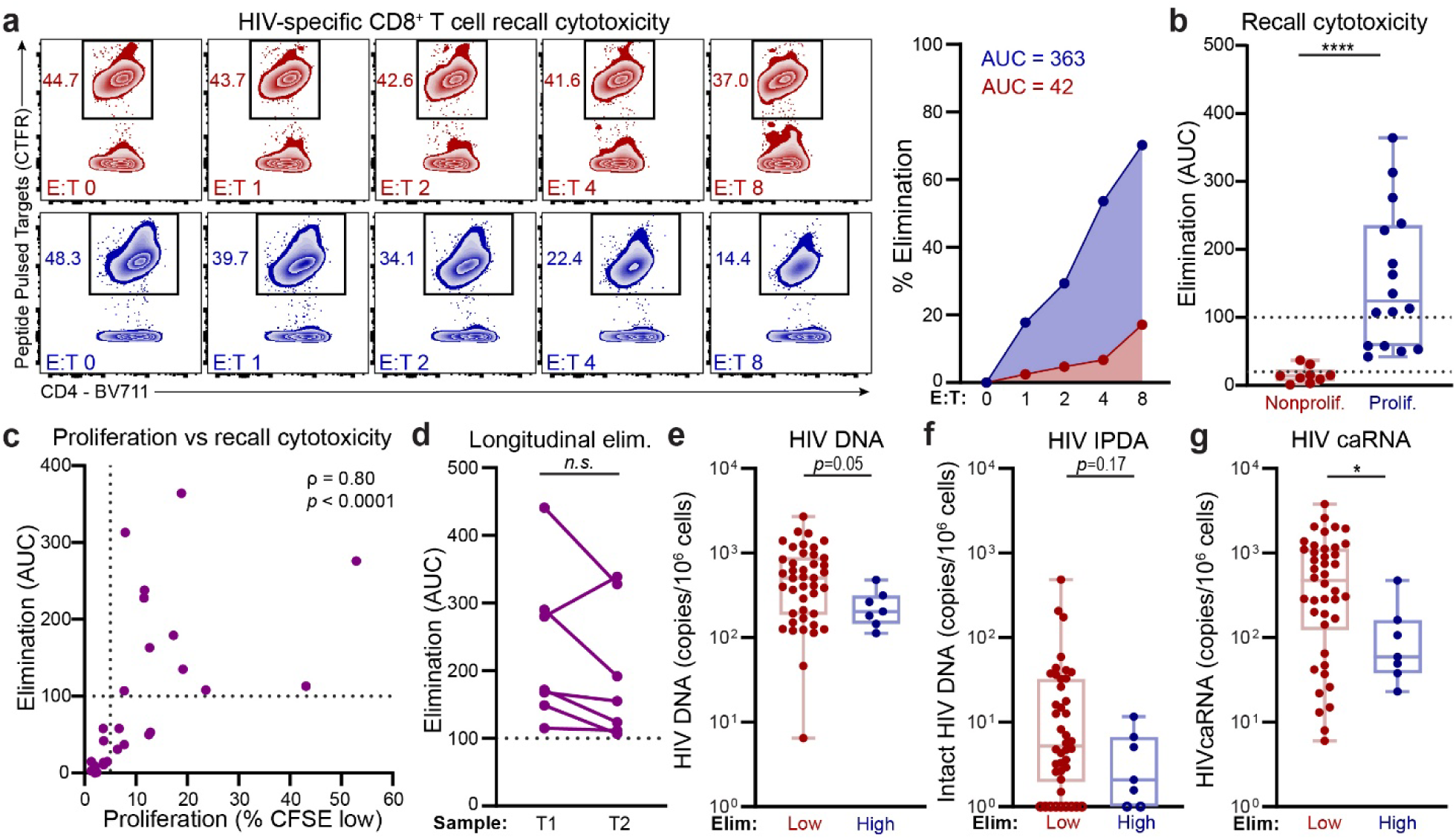
HIV-specific CD8^+^ T cell recall cytotoxicity is associated with smaller HIV-1 reservoirs. **(a)** Left: Example frequencies of residual Gag TPQDLNTML peptide-pulsed target CD4^+^ T cells labeled with CellTrace Far Red (CTFR) after coculture with autologous peptide-expanded effector CD8^+^ T cells at increasing effector:target (E:T) ratios for two responses with high (blue) or low (red) proliferative capacities. Right: Percent elimination plotted across E:T ratios for representative elimination responses shown on left, with area-under-curve (AUC) summary values. **(b)** Summary of recall cytotoxicities across all proliferative (% CFSE low ≥5%, *n*=16 responses from 14 participants) and a sampling of immunodominant nonproliferative (% CFSE low < 5%, *n*=9 responses from 8 participants) HIV-1 epitope-specific responses. Dashed lines at AUC=100 and AUC=20 represent thresholds for classification of responses with high, moderate, and low cytolytic capacities. \*\*\*\**p*<0.0001, Wilcoxon rank-sum test. **(c)** Spearman correlation (ρ) and corresponding correlation test of proliferative capacity measured by CFSE dilution assay versus recall cytotoxicity measured by expanded antigen-specific elimination assay (*n*=25 responses from 22 participants). Dashed lines at AUC=100 and 5% CFSE low represent thresholds for classification of responses with high cytolytic and proliferative capacities, respectively. **(d)** Recall cytotoxicity (elimination AUC) across longitudinal samples spanning 1-5 years in *n*=7 participants who had high recall cytotoxicity (AUC >100; *n.s.*, not significant; *p*=0.16, Wilcoxon matched-pairs signed rank test). **(e-g)** Measurements of total HIV-1 proviral DNA (e), intact HIV-1 proviral DNA measured by the intact HIV-1 provirus assay (IPDA, f), and cell-associated HIV-1 RNA (caRNA, g) copies per 10^6^ PBMCs from participants with HIV-specific CD8^+^ T cell responses of low (max. elim AUC <100, *n*=42 participants) or high (max. elim AUC >100, *n*=7 participants) cytolytic capacity. Open circles at 10^0^ represent values below the assay detection limit. \**p* < 0.05; *p-*values calculated by Wilcoxon rank-sum tests.

Using longitudinal specimens from seven participants with high recall cytotoxicity collected during continuous ART over a median 2.6 years (range 0.7-4.9 years), we assessed whether cytolytic potential was durably maintained. HIV-specific CD8^+^ T cell recall cytotoxicity remained high in all participants (Fig. 2d). In addition, proliferative CD8^+^ T cell responses from HEAL participants efficiently killed HIV-infected cells presenting naturally processed viral epitopes at levels comparable to those observed in spontaneous HIV-1 controllers (Extended Data Fig. 2). These results demonstrate that individuals with highly proliferative HIV-1 epitope-specific CD8^+^ T cell responses also durably maintain high cytotoxic recall potential against HIV-infected cells during ART and reveal an unexpectedly large minority (17%) of PWH harboring highly functional HIV-specific CD8^+^ T cell responses.

To investigate relationships between the functionality, frequency, and phenotype of virus-specific CD8^+^ T cells among PWH treated during chronic infection, we profiled immunodominant HIV-1 epitope-specific CD8^+^ T cells *ex vivo* using peptide-HLA (pHLA) tetramers. Although large responses were frequently nonproliferative (Fig. 1e), highly proliferative HIV-1 epitope-specific CD8^+^ T cells were typically present at higher *ex vivo* frequencies than nonproliferative cells (Extended Data Fig. 3a), suggesting that functional HIV-specific CD8^+^ T cells are also more expanded *in vivo*, consistent with their capacity to further expand and differentiate into cytotoxic effector cells *in vitro*. However, phenotypic analysis revealed no consistent distinguishing features of highly functional responses with respect to memory, activation, or differentiation states as defined by *ex vivo* staining for CD45RA, CD62L, CD38, PD-1, CD127, granzyme B, or TCF-1 (Extended Data Fig. 3b-g). These results indicate that superior CD8^+^ T cell responsiveness is not associated with reduced markers of immune activation or exhaustion and that phenotypic markers alone are insufficient to predict functional capacity during suppressive ART, when recent antigen exposure is limited.

### CD8^+^ T cell recall cytotoxicity is associated with reduced HIV-1 reservoir size and activity

To assess the relationship between highly cytolytic CD8^+^ T cells capable of mounting responses against autologous HIV-1 epitopes and persistent HIV-1 reservoirs during prolonged ART, we measured total and intact proviral DNA and unspliced cell-associated HIV-1 RNA (usRNA) in 49 participants with sustained virological suppression for at least one year. Participants with high CD8^+^ T cell proliferation and recall cytotoxicity harbored lower total (mean 85 vs. 254 copies/10^6^ PBMCs, Fig. 2e) and intact (mean 3.9 vs. 32.3 copies/10^6^ PBMCs) proviral HIV-1 DNA (Fig. 2f, Extended Data Fig. 4), and significantly lower cell-associated HIV-1 usRNA (mean 130 vs. 751 copies/10^6^ PBMCs, p=0.03, Fig. 2g) at the sample time points used for immunologic assessments.

These associations may indicate a role for CD8^+^ T cell recall cytotoxicity in limiting HIV-1 persistence, the converse, or both. To explore whether superior HIV-specific CD8^+^ T cell functionality during prolonged ART may associate with limited disease progression prior to ART, we analyzed 16 demographic, clinical, and treatment variables and found no significant associations with high cytolytic potential, including peak plasma viral load or nadir CD4 counts (Extended Data Fig. 5, Extended Data Table 1). These results argue against superior pre-ART viral control as the basis for enhanced CD8^+^ T cell function and support a potential role for recall cytotoxicity in limiting the size of transcriptionally active persistent HIV-1 reservoirs during prolonged ART.

### Peripheral immune responses to HIV-1 during analytical treatment interruption

We next evaluated HIV-1 rebound and *in vivo* cellular immune responses during analytical treatment interruption (ATI) in one HEAL participant (H047). Plasma viral loads were measured weekly and PBMCs were obtained bi-weekly during ATI (Extended Data Table 2). Plasma viremia became detectable by two weeks post-ATI and peaked at 5.16 log_10_ HIV-1 RNA copies/ml at week 5 post-ATI, followed by a spontaneous 3.9-fold reduction over 6 days to 4.56 log_10_ copies/ml by week 6 post-ATI (Fig. 3a). Per protocol-defined criteria, ART was restarted after collection of the week 6 blood sample and plasma viremia re-suppressed to undetectable levels (<20 copies/mL) within two weeks (Fig. 3a, Extended Data Table 2). HIV-1 cell-associated RNA (caRNA) levels increased during ATI but not to levels commensurate with plasma viremia; caRNA returned to pre-ATI levels 4 weeks after ART re-initiation and remained unchanged one year later (Extended Data Fig. 6). Unspliced HIV-1 RNA exceeded polyadenylated HIV-1 RNA levels during ART, consistent with prior reports^34^, but this relationship reversed during detectable viremia. One year after ATI, HIV-1 persistence markers were similar to pre-ATI levels.

**Fig. 3.**
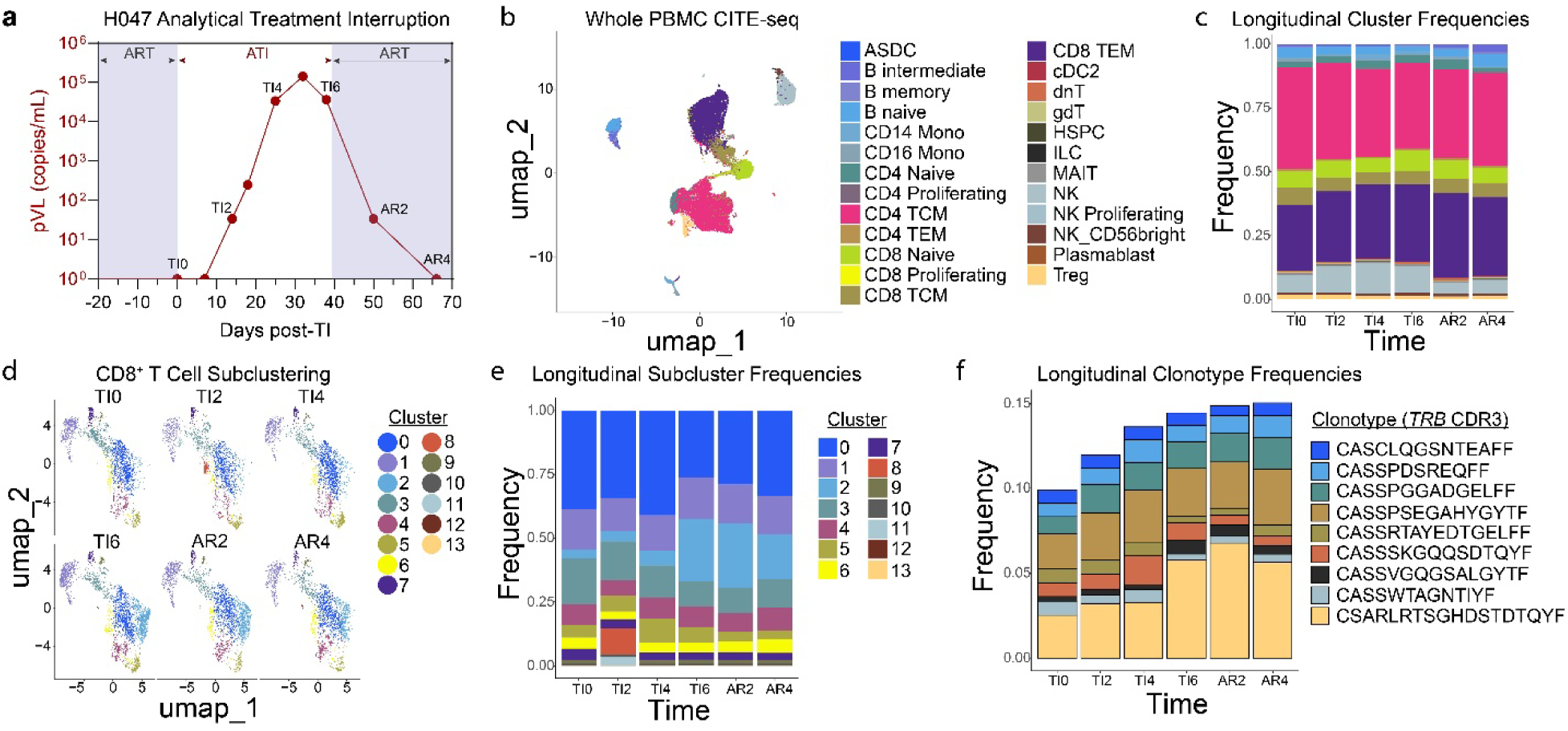
Interruption of antiretroviral therapy initiates sequential NK and clonal CD8^+^ T cell expansions. **(a)** Longitudinal plasma HIV-1 viral loads (pVL, log_10_ scale) monitored during ATI in H047. Unshaded region indicates beginning of ATI at day 0 (TI0) and resumption of ART following the sample collected on day 38 (TI6). Biweekly PBMC sample collections are labeled; TI, treatment interruption; AR, ART-restart; numbers represent weeks. **(b)** WNN analysis of a 5’ CITE-seq dataset of PBMC and 154 surface proteins, displayed as a UMAP visualization. ASDC, AXL^⁺^ SIGLEC6^⁺^ dendritic cells; B intermediate, transitional/intermediate stage B cells; CD14 Mono, CD14^⁺^ classical monocytes; CD16 Mono, CD16^⁺^ nonclassical monocytes; TCM, central memory T cells; TEM, effector-memory T cells; cDC2, type-2 conventional dendritic cells; dnT, double-negative T cells (CD4^⁻^CD8^⁻^); gdT, γδ T cells; HSPC, hematopoietic stem and progenitor cells; ILC, innate lymphoid cells; MAIT, mucosal-associated invariant T cells; Treg, regulatory T cells. **(c)** Stacked bar plot of PBMC proportions over time **(d)** Longitudinal CD8^+^ T cell sub-clustering at resolution 0.2 identifies expansion of cluster 2, most notable at TI6, that begins to contract by AR4. **(e)** Stacked bar plots of CD8^+^ T cell subcluster frequencies from d at each ATI sample timepoint. **(f)** Stacked bar plots of CD8^+^ T cell clonotype frequencies, as defined by complementary-determining region 3 (CDR3) sequence of the T cell receptor beta (*TRB*) chain and plotted as fractions of total CD8^+^ T cells.

To comprehensively profile peripheral immune responses during and after ATI, we performed integrated single-cell multiomics analyses on PBMC collected across six time points and assessed changes in T cell receptor (TCR) and B cell receptor (BCR) clonality, surface protein and gene expression. A total of 37,462 PBMC across six time points were included in longitudinal CITE-seq and immune profiling analyses (Fig. 3b, Supplementary Data 2). While the frequencies of most immune cell populations remained stable during treatment interruption, we observed a 1.7-1.9-fold expansion of NK cells that began in week 2 of ATI, remained elevated through week 6 of ATI, and then contracted to approximately half of pre-ATI levels after ART was restarted (Fig. 3c). This NK cell expansion was characterized by a cytolytic terminal effector phenotype (Extended Data Fig. 7)^35, 36^. A sub-clustering analysis of CD8^+^ T cells revealed a 7-8-fold expansion of one subcluster (cluster 2) that began in week 4 of ATI, persisted through ART restart, and then contracted 4 weeks after ART re-initiation (Fig. 3d-e, Extended Data Fig. 8a).

Dynamic shifts in TCR clonotypes accompanied ATI (Fig. 3f). The most prevalent CD8 clonotype pre-ATI comprised 2.7% of all clonotypes (CSARLRTSGHDSTDTQYF). This and a sub-dominant clonotype (CASSVGQGSALGYTF) each expanded 2.7-2.9-fold during ATI and were enriched among cluster 2, whereas other minor clonotypes decreased up to 2.5-fold in frequency (Supplementary Data 2). A transient CD8^+^ T cell population (cluster 8) was identified in the second week of treatment interruption when viremia was first detected (Fig. 3d-e). This cluster expressed TCRs matching cluster 2 (Supplementary Data 2) but was distinguished by a hypoxia-adapted, glycolysis-driven cellular program (Extended Data Fig. 8b-f). Modest changes in B cell subsets during ATI were not associated with detectable clonal expansion, while changes in CD4^+^ T cell subpopulation proportions were not observed (Extended Data Fig. 9). Together, these findings reveal the detailed kinetics of concurrent and sequential responses to HIV-1 recrudescence across immune cell subsets.

### *In vivo* CD8^+^ T cell recall cytotoxicity coincides with attenuated rebound viremia

Prior to ATI, the immunodominant CD8^+^ T cell response in participant H047 targeted the HLA-B*44-restricted Gag AEQASQEVKNW (AW11) epitope and comprised approximately 4.5% of total CD8^+^ T cells (Fig. 4a). At baseline, this response was highly proliferative (12% CFSE-low) and moderately cytolytic (52 AUC) *in vitro* (Fig. 4a). Although the presence of this large, functional HIV-specific CD8^+^ T cell population did not prevent virus rebound, rapid attenuation of viremia observed at 6 weeks post-ATI prior to ART reinitiation was associated with concurrent *in vivo* expansion of Gag AW11-specific cells from 4.5% to 11.4% of total CD8^+^ T cells (Fig. 4b). In contrast, a subdominant HLA-B*18-restricted Nef YPLTFGWCF (YF9)-specific CD8^+^ T cell response did not expand during ATI (Fig. 4b). The observed 2.5-fold *in vivo* expansion of AW11-specific cells was remarkably similar in magnitude to the 2.7-fold expansion of these cells following *in vitro* peptide stimulation (Fig. 4a), supporting the predictive value of *in vitro* proliferation assays in modeling the HIV-specific CD8^+^ T cell response to virus reemergence. One year following ATI, the AW11-specific CD8^+^ T cell response retained comparable *ex vivo* frequency, *in vitro* proliferative capacity and recall cytotoxicity to pre-ATI (Fig. 4c), suggesting that ATI did not have a detrimental impact on the quality of HIV-specific CD8^+^ T cell immunity in this participant.

**Fig. 4:**
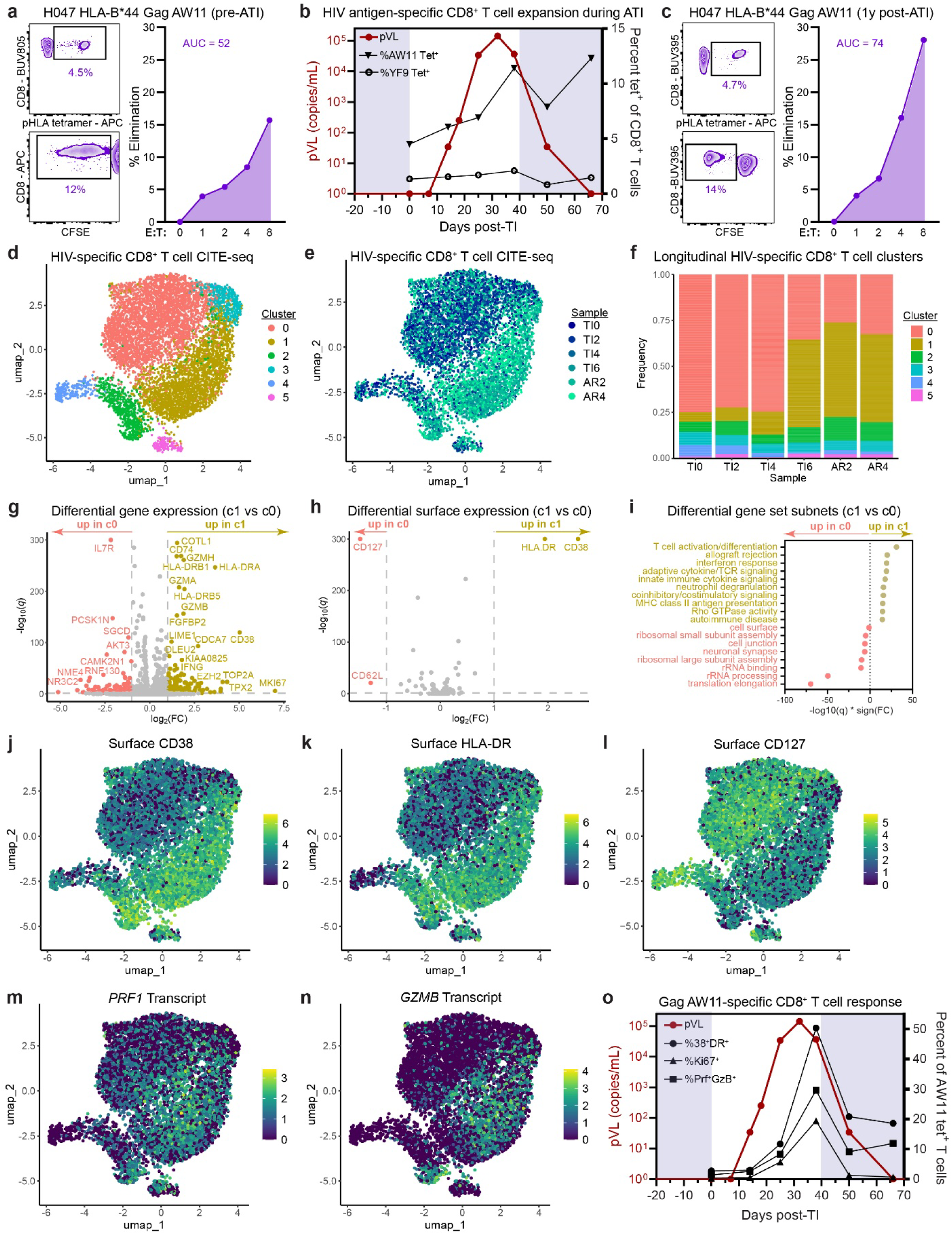
*In vivo* HIV-specific CD8^+^ T cell expansion and cytolytic effector differentiation accompany attenuated rebound viremia during ATI. **(a)** Summary of HLA-B*44:02-restricted HIV-1 Gag AEQASQEVKNW (AW11) CD8^+^ T cell response frequency (top left), proliferative capacity (bottom left), and secondary cytolytic potential (right) in participant H047 prior to analytical treatment interruption (ATI). **(b)** Longitudinal plasma HIV-1 viral loads (pVL; left axis, log_10_ scale) monitored during ATI in H047. Unshaded region indicates beginning of ATI at day 0 and resumption of ART following the sample collected on day 38 (red circles). Immunodominant AW11-specific (black triangles), and subdominant Nef YF9-specific CD8^+^ T cell frequencies measured by pHLA tetramer (Tet) staining during ATI in H047 (right axis, linear scale). **(c)** Summary of AW11-specific CD8^+^ T cell response frequency (top left), proliferative capacity (bottom left), and secondary cytolytic potential (right) in participant H047 one year post-ATI. **(d-e)** UMAP projection of single-cell transcriptomes derived from AW11-specific CD8^+^ T cells during ATI in participant H047, colored by shared nearest neighbor clustering (d) or by sample time-point (e). **(f)** Frequencies of clusters shown in d at each sample time-point during ATI. **(g-h)** Summary of differential gene (g) and surface marker (h) expression between clusters 0 and 1 among AW11-specific CD8^+^ T cells. Dashed lines represent significance (*q*<0.05) and fold change (FC>2) thresholds. **(i)** Summary of the top significantly differential gene set subnets between cluster 0 and cluster 1 derived from gene set network analysis (GSNA). **(j-n)** UMAP projection of single-cell transcriptomes derived from AW11-specific CD8^+^ T cells during ATI in H047, colored by log-normalized expression of surface CD38 (j), HLA-DR (k), CD127 (l), *PRF1* transcript (m), or *GZMB* transcript (n). **(o)** Longitudinal HIV-1 pVL (red circles, left axis, log_10_ scale), *in vivo* HIV-specific CD8^+^ T cell activation measured by CD38 and HLA-DR co-expression (black circles, right axis, linear scale), proliferation measured by Ki-67 (black triangles, right axis, linear scale), and cytotoxic effector differentiation measured by perforin and granzyme B coexpression (black squares, right axis, linear scale) by flow cytometry among AW11 tet^+^ CD8^+^ T cells during ATI in H047.

To further evaluate the CD8^+^ T cell response to HIV-1 recrudescence at epitope-specific, temporal, and single-cell resolution, we performed CITE-seq on 7,500 Gag AW11-specific CD8^+^ T cells isolated from six sample time points spanning ATI. These cells segregated into six clusters based upon differential gene expression (Fig. 4d), of which cluster 0 predominated at early time points (Fig. 4e,f). After re-emergence of viremia, the frequency of cells in cluster 0 decreased and a majority of HIV-specific CD8^+^ T cells were instead found in cluster 1 (Fig. 4e,f). This transition was associated with an increase in expression of genes and surface proteins associated with T cell activation and cytotoxic effector differentiation (Fig. 4g-i, Supplementary Data 3), including CD38, HLA-DR, *PRF1*, and *GZMB*; and a concomitant decrease in T cell quiescence as indicated by *IL7R*/CD127 (Fig. 4j-n). Most HIV-specific cells remained in this differentiated state for at least four weeks following reinitiation of ART as viremia declined (Fig. 4f). To validate these findings by orthogonal methods, we also assayed longitudinal H047 ATI samples by flow cytometry and assessed *in vivo* HIV-specific CD8^+^ T cell activation as measured by CD38 and HLA-DR co-expression, proliferation as measured by Ki-67 staining, and secondary cytotoxic effector differentiation as measured by perforin and granzyme B co-expression among pHLA multimer^+^ cells. Rapid reduction of plasma viral load was accompanied by pronounced increases in activation, proliferation, and cytotoxicity of HIV-specific CD8^+^ T cells (Fig. 4o). These results provide new insights into the dynamics of a functional HIV-specific CD8^+^ T cell response to the resurgence of persistent virus and suggest that recall cytotoxicity from a resting memory state may contribute to attenuation, but not prevention, of rebound viremia.

### CD8^+^ T cell clonotypes with distinct phenotypes expand in parallel during ATI

Next, to better understand the clonalities and phenotypes of T cells responding to HIV-1 rebound in H047, we performed clonotypic analysis based on TCRβ CDR3 sequences. The Gag AW11-specific response in H047 was oligoclonal, with more than 75% of cells belonging to a single highly expanded clonotype (Fig. 5a). AW11-specific clonotypes represented the 8 most abundant TCRs among total CD8^+^ T cells identified in our single-cell PBMC analysis, including those in CD8^+^ T cell subclusters 2 and 8 that expanded following rebound viremia (Fig. 3f, Supplementary Data 2), further validating our epitope mapping approach and underscoring the presence of remarkably large epitope-specific clones. As the AW11-specific CD8^+^ T cell population expanded in response to HIV-1 rebound (Fig. 4b), its clonotypic composition was largely maintained, except for one minor clonotype (CASSLYSNQPQHF) that expanded from less than one percent to greater than four percent of the response (Fig. 5b). Clonotypes differed significantly in gene and surface expression at baseline (Fig. 5c,d), with the dominant clonotype comprising clusters 0, 1, 3, and 5, and subdominant clonotypes comprising clusters 2 and 4 (Fig. 5a).

**Fig. 5:**
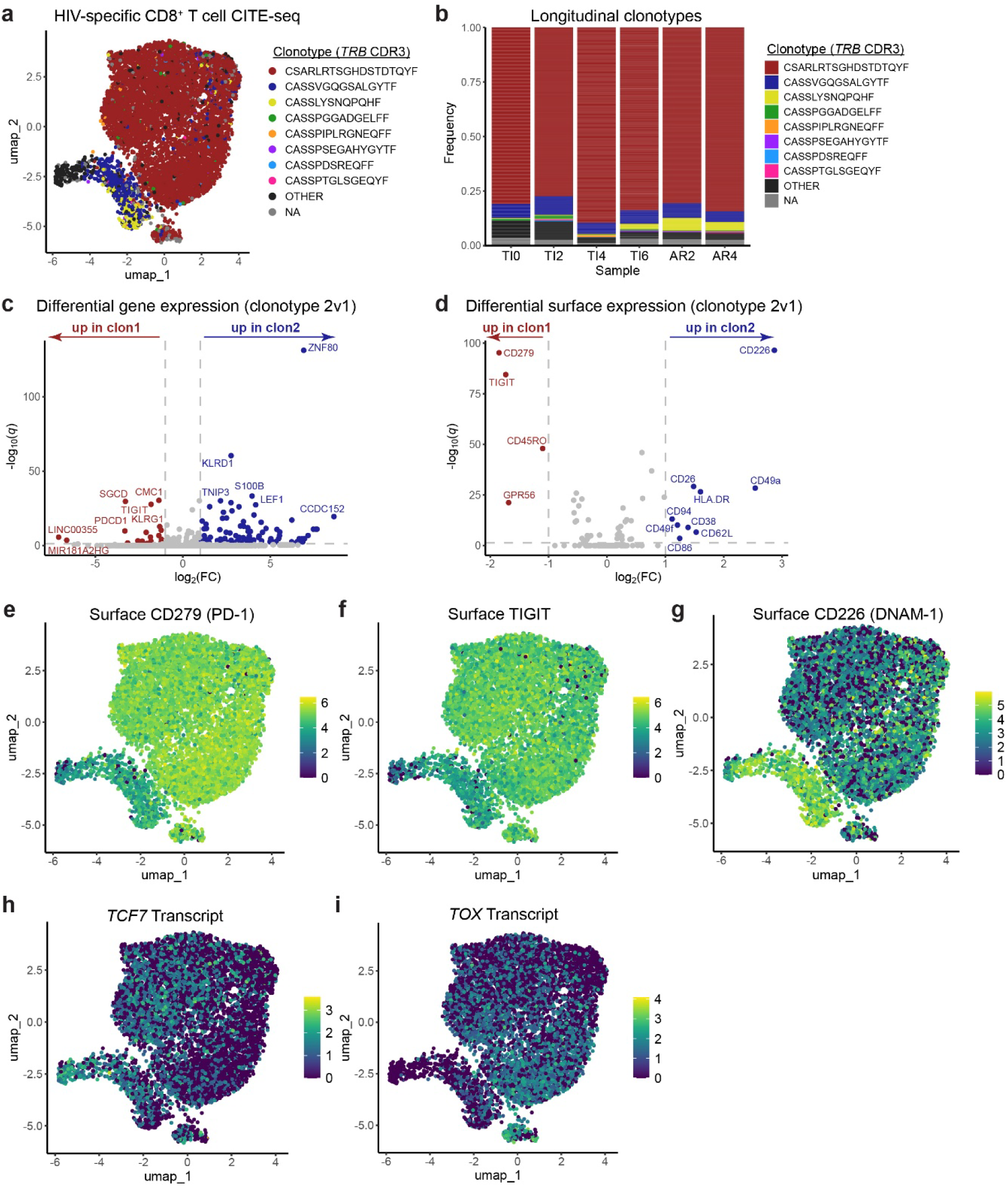
T cell clonotypes with progenitor-exhausted and quiescent memory markers expand in parallel during antiretroviral therapy interruption. **(a)** UMAP projection of single-cell transcriptomes derived from AW11-specific CD8^+^ T cells during ATI in H047, colored by TCR clonotype and labeled by *TRB* CDR3 sequences. **(b)** Frequencies of clonotypes shown in A at each sample time-point during ATI. **(c-d)** Summary of differential gene (c) and surface marker (d) expression between clonotypes CSARLRTSGHDSTDTQYF (clon1) and CASSVGQGSALGYTF (clon2) among AW11-specific CD8^+^ T cells. Dashed lines represent significance (*q*<0.05) and fold change (FC>2) thresholds. **(e-l)** UMAP projection of single-cell transcriptomes derived from AW11-specific CD8^+^ T cells during ATI in H047, colored by log-normalized expression of surface CD279/PD-1 (e), TIGIT (f), CD226 (g), *TCF7* transcript (h), or *TOX* transcript (i).

Prior to rebound viremia, the dominant clonotype expressed T cell exhaustion gene and surface signatures, marked by elevated PD-1, TIGIT, and *TOX*, whereas the largest subdominant clonotype appeared more quiescent with elevated expression of CD226 and *TCF7* (Fig. 5c-i). Despite their phenotypic differences, both clonotypes expanded similarly by 2.7-2.9 fold *in vivo* during ATI (Supplementary Data 2), maintained their relative frequencies within AW11-specific CD8^+^ T cells during their expansion in response to recrudescent viremia (Figs. 5b, 4b), and demonstrated comparable increases in activation and cytotoxic effector differentiation (Fig. 4d-e,j-n). Notably, the *TCF7*^+^ *TOX*^+^ PD-1^+^ CD8^+^ T cells that comprised the dominant clonotype at baseline (cluster 0) resembled progenitor-exhausted cells that have been previously shown to be important for control of chronic viral infections and for memory formation upon removal of persistent antigen^37, 38, 39, 40, 41^, suggesting that this may also occur during ART-mediated HIV-1 suppression. The lack of an observed expansion disadvantage of this progenitor-exhausted clonotype 1 relative to the more quiescent and less exhausted clonotype 2 indicates their comparable functionality *in vivo*, consistent with our *in vitro* phenotyping results (Extended Data Fig. 3). Our collective results suggest that irrespective of phenotypic markers, CD8^+^ T cells with strong proliferative and cytolytic capacities may cooperatively limit virus replication in a subset of PWH treated during chronic infection.

## DISCUSSION

In this study, we investigated the prevalence of functional CD8^+^ T cell memory responses targeting autologous HIV-1 epitopes during prolonged ART and their association with viral persistence markers. We observed unexpectedly high HIV-specific recall cytotoxicity in 17% of PWH who initiated ART during chronic progressive infection. These individuals harbored smaller and less transcriptionally active persistent viral reservoirs relative to individuals with poor HIV-specific recall cytotoxicity. We also observed *in vivo* expansion and recall cytotoxicity of HIV-1 epitope-specific memory CD8^+^ T cells concurrently with attenuated rebound viremia in a participant who underwent ATI. We thoroughly characterized this epitope-specific CD8^+^ T cell response to recrudescent viremia at temporal, clonotypic, and single-cell resolution, revealing that clonotypes with progenitor-exhausted and quiescent memory-like features expand comparably *in vivo* during attenuated viremia following ATI. Together, these findings reveal the prevalence of functional memory CD8^+^ T cells targeting autologous HIV-1 during chronic treated infection and suggest their potential importance in limiting HIV-1 persistence and rebound viremia, offering valuable guidance for the development of combination therapies to achieve durable ART-free remission.

Two related goals for HIV-1 cure approaches are to eliminate persistent virus reservoirs and elicit durable immune control to obviate the need for lifelong ART. CD8^+^ T cells may contribute substantively toward achieving both goals. It is well established that CD8^+^ T cell proliferation and recall cytotoxicity are key features of spontaneous HIV-1 control^2, 3, 4, 5, 6^ and that these functions are generally impaired in noncontrollers, even after suppressive ART^21^. We and others recently reported that CD8^+^ T cell proliferation and recall cytotoxicity are also distinguishing features of post-intervention HIV-1 control elicited by immunotherapies in ATI trials^23, 24, 25^, underscoring the existence of cells with these functional properties in some proportion of noncontrollers on ART and their importance to cure approaches. There have been limited prior reports of CD8^+^ T cell functional restoration after long-term ART in PWH treated during chronic infection^22, 42^, but the prevalence and impact on HIV-1 persistence and rebound viremia were not clear, prompting the present study. We found that the prevalence of CD8^+^ T cells with high recall cytotoxicity among PWH on prolonged suppressive ART initiated during chronic HIV infection was unexpectedly high at 17%, which is more than 50 times the rate of spontaneous HIV-1 control^43^ and more than 15 times the rate of post-treatment control in non-interventional ATI studies of people treated during chronic infection^44^. Intriguingly, the prevalence of these functional responses was comparable to the frequency of durable post-intervention control induced by bNAbs^26, 27, 28, 29^, consistent with our recent report that CD8^+^ T cell proliferation and recall cytotoxicity precede this outcome^23^. Strategies to elicit these functions in the remaining majority of PWH may enable more broadly successful HIV-1 cure approaches.

The relationship between HIV-specific CD8^+^ T cell functionality and HIV-1 persistence during prolonged ART in noncontrollers is not well understood. Prior studies have inferred immune selection of virus reservoirs by HIV-1 sequencing analysis^45, 46, 47, 48^ but have not demonstrated direct associations with immune cell functionality. In our study, participants with high recall cytotoxicity against autologous HIV-1 epitopes harbored smaller and less transcriptionally active reservoirs. It is plausible that these associations could represent effects of functional CD8^+^ T cells on HIV-1 persistence, the converse, or both. Prior studies have demonstrated that CD8^+^ T cells contribute to viral suppression during ART^49^ and that early ART initiation is associated with smaller HIV-1 reservoirs and preserved CD8^+^ T cell functionality^7, 8, 50^. Repeated ART interruption or spontaneous HIV-1 expression during ART can also impact CD8^+^ T cell features^51, 52, 53, 54, 55^. Importantly, neither treatment lapses, earlier ART initiation, prior spontaneous control, nor limited disease progression were evident among the 17% of participants with high HIV-specific recall cytotoxicity. While viral transcription is a prerequisite for antigen presentation to CD8^+^ T cells, intactness is not^56^. Thus, it is noteworthy that CD8^+^ T cell functionality was most strongly associated with the size of the transcriptionally active portion of the reservoir and most weakly associated with the size of the intact portion of the reservoir, further suggesting an impact of functional CD8^+^ T cells on HIV-1 persistence during ART. A recent study from an independent cohort of PWH reported functional rejuvenation following long-term ART that was associated with clonal succession^22^. The dominant clonotype functionally responding to treatment interruption in our study exhibited *ex vivo* signatures of chronic antigen stimulation, which may be inconsistent with recent clonal succession. Further research is warranted to identify precise mechanisms by which CD8^+^ T cell functionality can be maintained or restored in PWH. While participants with superior HIV-specific CD8^+^ T cell functionality had smaller virus reservoirs, it is important to note that such functionality was insufficient for HIV-1 eradication and that several participants with poor functionality also harbored small reservoirs. These results prompt the evaluation of strategies to boost immune-mediated reservoir clearance, which may inform the development of multimodal therapies to elicit durable HIV-1 remission.

Persistent HIV-1 reservoirs are sensitive to recognition by a subset of circulating T cells^57, 58^, yet most HIV-specific CD8^+^ T cells exhibit profound dysfunction associated with exhaustion-associated inhibitory receptors such as PD-1^59^. Although suppressive ART reduces inhibitory receptor expression, it typically fails to restore HIV-specific CD8^+^ T cell proliferation and cytotoxicity^21^. Consistent with this, we observed similar levels of PD-1 expression on functional and dysfunctional HIV-specific CD8^+^ T cells *ex vivo* during prolonged ART and similar *in vivo* expansion of PD-1^high^ and PD-1^low^ HIV-specific CD8^+^ T cells during ATI, further establishing a disconnect between inhibitory receptor expression and CD8^+^ T cell functionality. Similarly, response functionality in this cohort was not significantly associated with *ex vivo* memory phenotypes. Together, these findings demonstrate that phenotypic state alone fails to predict antiviral recall capacity and highlight the need for functional profiling to define immune potential in treated HIV-1 infection, especially in settings in which prolonged ART-mediated suppression of HIV-1 limits *in vivo* antigen exposure. In addition, because mutational escape from CD8^+^ T cell recognition can result in retention of functionality through viral evasion of chronic stimulation^32^, evaluation of CD8^+^ T cell function at epitope-specific resolution and following re-exposure to autologous cognate antigen was critical to accurately determining the prevalence of functional CD8^+^ T cells in this cohort. Five of the sixteen proliferative responses we identified did not recognize autologous epitopes and another eight recognized autologous epitopes that differed from HIV-1 consensus sequences. Thus, absent evaluation of autologous cross-recognition, the five escaped responses could have been incorrectly interpreted as functional, or half of the functional responses could be incorrectly assumed to have escaped recognition. For these reasons, commonly used biomarkers and functionality assessments involving stimulation with consensus viral peptide pools or laboratory virus strains may inaccurately estimate CD8^+^ T cell function.

Recall cytotoxicity of a large, functional HIV epitope-specific CD8^+^ T cell response in one participant lagged rebound viremia, failing to prevent it. This result suggests that CD8^+^ T cells capable of mounting cytotoxic recall responses may be insufficient to prevent virus rebound. As memory CD8^+^ T cells generally downregulate cytotoxic effector proteins in the absence of recent antigen exposure, they may be unlikely to prevent HIV-1 rebound without additional interventions and, consistent with this, are generally not associated with time to onset of viral rebound, which itself is not well correlated with set-point plasma viral load^60, 61^. However, CD8^+^ T cell recall cytotoxicity may play important roles in limiting, suppressing, or controlling recrudescent viremia. Due to clinical concerns, most ATI studies re-initiate ART before the ability of secondary effector cell responses to control rebound viremia can be ascertained^62^. Although limited to a single participant, our study was no exception; ART restart criteria were met before set-point viremia was reached. Nevertheless, viremia was blunted concurrently with the *in vivo* expansion of this response prior to ART reinitiation, suggesting a potential role in limiting the extent of HIV-1 replication. Recent evidence suggests that the addition of exogenous bNAbs at the time of ATI may help to provide a head-start for such recall responses, allowing for durable prevention of virus rebound^23, 25^, and that therapeutic vaccination to boost CD8^+^ T cell immunity may further accelerate this recall response at the time of virus recrudescence, enabling superior post-rebound control^24, 63^.

Our study has limitations. The absence of matched longitudinal specimens prior to ART initiation precluded definitive inference regarding the mechanisms by which CD8^+^ T cell functionality is preserved or restored during prolonged therapy. Analyses of immune dynamics during ATI were limited to a single participant, and viremia could not be followed beyond week 6 post-ATI due to protocol-defined ART restart criteria. Accordingly, insights from this ATI should be interpreted in the context of the broader literature and used to inform future interventional studies designed to prevent or control HIV-1 rebound. Our analyses were restricted to peripheral blood, as tissue samples from key anatomical sites of HIV-1 persistence and recrudescence were not available. Despite these limitations, our results demonstrate robust CD8^+^ T cell recall cytotoxicity in a proportion of people with HIV who initiated treatment during chronic infection, the largest population in need of an effective and durable HIV-1 cure. Our findings highlight recall cytotoxicity of memory CD8^+^ T cells targeting autologous HIV-1 as an important parameter in the development and evaluation of multimodal immunotherapeutic strategies to combat HIV-1 persistence. Efforts to prime CD8^+^ T cells to more rapidly contain and control viral recrudescence at the time of ART interruption, in concert with latency reversal and/or orthogonal suppressive modalities, may be required to achieve durable HIV-1 remission.

## METHODS

### Study design

This non-interventional study was designed to assess the range of cytolytic activity among CD8^+^ T cells targeting autologous HIV-1 epitopes during prolonged ART and its potential impact on virus persistence. Peripheral blood was collected from ART-suppressed PWH enrolled in the HIV Eradication and Latency (HEAL) cohort, a longitudinal study of PWH. The institutional review board at Brigham and Women’s Hospital (Mass General Brigham Human Research Committee – MGB HRC) approved the study protocol. Written informed consent was obtained from all participants. Experimenters were blinded to all clinical data. Cohort size (*n*= 60) was determined by specimen availability. Plasma HIV-1 viral loads, complete blood counts, and demographic information were extracted from electronic medical records with participant consent. All participants were treated with suppressive ART for a minimum of one year (median 19 years, IQR 11-22 years) prior to sample collection. PBMCs were isolated by density gradient centrifugation and cryopreserved. PBMCs from a separate group of spontaneous HIV-1 controllers who maintained plasma viral loads below 2,000 HIV-1 RNA copies/ml for several years without ART from the Ragon Institute cohort were included for comparison to HEAL participants across immunologic assays following MGB HRC secondary use approval.

Participant H047 enrolled in an elective, IRB-approved HEAL ATI substudy after meeting stringent enrollment criteria derived from consensus recommendations for ATI studies^62^, risk counseling, and written informed consent. Peripheral blood was collected bi-weekly and plasma viral load was monitored weekly during ATI. Pre-determined ART restart criteria included request by the participant or their HIV-1 healthcare provider, pregnancy, symptomatic HIV-1 disease, confirmed absolute CD4 value <350 cells/µl or CD4% less than 15, plasma viral load greater than or equal to 1,000 HIV-1 RNA copies/ml for 4 weeks or greater than 100,000 copies/ml at any time. After ART reinitiation, plasma viral loads and CD4 counts were monitored to ensure full virus re-suppression was achieved.

### HLA genotyping

Frozen dry cell pellets were sent to the American Red Cross Histocompatibility Laboratory Services (Dedham, Mass.) for high-resolution genotyping of the *HLA-A, -B,* and *-C* loci. *HLA-B* alleles associated with low or high plasma viral loads in a genome-wide association study of natural HIV-1 infection^31^ were considered as protective (*HLA-B*14, B*27, B*52, B*57*) or risk (*HLA-B*07, B*08, B*35*), respectively, while all other *HLA-*B alleles were considered neutral. Allele frequencies were compared to the United States population^30^ and to 10 spontaneous HIV-1 controllers who were genotyped previously by the Carrington laboratory at the United States National Cancer Institute.

### HIV-1 epitope-specific response mapping by IFN-**γ** elispot

Frozen PBMCs were thawed at 37°C, recovered in RPMI media (Sigma-Aldrich) supplemented with 10% fetal bovine serum (FBS, Sigma; R10) overnight, resuspended at 1x10^6^/mL in R10, and plated 200 µL per well in Immobilon-P 96-well microtiter plates (Millipore) pre-coated with 2 µg/mL anti-IFN-γ (clone DK1, Mabtech). Individual HLA-optimal HIV-1 peptides matched to each subject’s *HLA* genotype, as listed previously^6^, were added at 1 µM and incubated at 37°C overnight. Triplicate negative control wells did not receive peptide and positive control wells were treated with 1 µg/ml anti-CD3 (clone OKT3, Biolegend) and 1 µg/ml anti-CD28 (clone CD28.8, Biolegend) antibodies. ELISPOT assay was performed using manufacturer’s protocol with anti-IFN-γ (clone 1-DK1, Mabtech) capture, biotinylated anti-IFN-γ (clone B6-1, Mabtech) detection, Streptavidin-ALP (Mabtech) and AP Conjugated Substrate (BioRad) followed by disinfection with 0.05% Tween-20 (Thermo Fisher) and analysis using S6 Macro Analyzer (CTL Analyzers). Responses greater than 10 spots per well (50 spots per 10^6^ PBMCs) and 3-fold above negative controls were scored as positive. Of 66 participants screened, 6 (9%) were excluded from downstream analyses due to low cell viability and/or high nonspecific background in elispot, which precluded interpretation of results.

### Proliferation assay

Proliferation was measured by CFSE dilution as described previously^6^. Briefly, frozen PBMCs were thawed at 37°C, recovered in RPMI media (Sigma-Aldrich) supplemented with 10% fetal bovine serum (FBS, Sigma; R10) overnight, then stained at 37°C for 20 minutes with 0.5 µM CellTrace CFSE (Thermo Fisher) as per manufacturer’s protocol. Cells were then quenched and washed twice with R10 media, resuspended at 1x10^6^/mL in R10, and plated at 200 µL per well in 96-well round-bottom polystyrene plates (Corning). Individual HLA-optimal HIV-1 peptides matched to each subject’s *HLA* genotype, as listed previously^6^, were added at 1 µM and incubated at 37°C for 6 days before flow cytometric assessment. Triplicate negative control wells did not receive peptide and positive control wells received 1 µg/ml anti-CD3 (clone OKT3, Biolegend) and 1 µg/ml anti-CD28 (clone CD28.8, Biolegend) antibodies. On day 6, cells were stained using Live/Dead Violet viability dye (Thermo Fisher), AlexaFluor700-anti-CD3 (clone SK7, Biolegend), and APC-anti-CD8 (clone RPA-T8, Biolegend), then analyzed by flow cytometry. Responses above 5% CFSE-low, equivalent to the mean proliferation of HIV-specific CD8^+^ T cell responses in spontaneous controllers, were considered to have high proliferative capacity and were subsequently validated in triplicate; reported values represent averages of triplicates.

### HIV-1 proviral epitope sequencing

Genomic DNA was extracted from CD4^+^ T cells isolated from 20 million PBMCs per sample using the EasySep CD4^+^ T cell isolation kit (StemCell Technologies) followed by the QIAamp DNA Blood Mini kit (Qiagen) via the manufacturer’s protocols. PCR amplification of HIV-1 proviral DNA was performed using nested-PCR primers specific for *gag*, *pol*, and/or 3’ half amplicons, as described previously^64^. Technical replicates for each PCR amplicon of correct size were gel-purified (Qiagen) and pooled for sequencing. Sequencing via Illumina MiSeq, assembly, and alignment to HXB2 reference were performed by the Mass General Brigham Center for Computational & Integrative Biology DNA Core (Boston, Mass) for amplicons containing epitopes targeted by HIV-specific CD8^+^ T cell responses of interest as determined by functional response mapping experiments. In cases where multiple autologous epitope variants were detected, all were tested for cross-recognition.

### Variant peptide cross-recognition assay

HIV-1 epitope-specific responses for which epitope variants were identified among autologous provirus were tested for cross-recognition of or mutational escape from CD8^+^ T cell recognition. PBMCs were stimulated for 4 hours with 1 µM of HLA-optimal peptides of clade B HIV-1 consensus and each autologous sequence observed in provirus. BV711-conjugated anti-CD107A (clone H4A3, Biolegend) was included during stimulation to measure degranulation. GolgiStop and GolgiPlug (BD Biosciences) were added 2 hours post-stimulation to enable intracellular cytokine staining. Cells were stained with Live/Dead Violet, BV605-conjugated anti-CD3 (clone SK7, Biolegend) and BUV395-conjugated anti-CD8 (clone RPA-T8, BD Biosciences), fixed and permeabilized using Cytofix/Cytoperm (BD Biosciences), stained for intracellular PE-Cy7-conjugated anti-IFN-γ (clone B27, Biolegend) and analyzed via flow cytometry. Recognition was considered maintained if the frequency of CD107A^+^ IFN-γ^+^ CD8^+^ T cells upon autologous variant peptide stimulation was greater than 50% for all observed variants or escaped if less than 50% that of clade B consensus peptide stimulation for any observed variant.

### Phenotypic cytometry

Peptide-HLA monomers for responses of interest were purchased from ImmunAware (Copenhagen, Denmark). Tetramers were produced by multimerization with APC-conjugated streptavidin (Biolegend) as per manufacturer’s protocol and stored at 4°C for a maximum of 24 weeks prior to use. Staining was performed using 4 nM individual APC-conjugated pHLA tetramers at 4°C for 30 minutes after 30-minute pre-treatment with 50 nM dasatinib to prevent *in vitro* cell activation and activation-induced cell death for phenotypic analysis. Cells were then stained with Live/Dead Near-IR viability dye (Thermo Fisher), BV510-conjugated anti-CD3 (clone SK7, Biolegend), BUV805-anti-CD8 (clone RPA-T8, BD Biosciences), BUV395-anti-CD45RA (clone HI100, BD Biosciences, RB780-anti-CD62L (clone DREG-56, BD Biosciences), BV711-anti-CD127 (clone A019D5, Biolegend), PE-Dazzle 594-anti-CD38 (clone HB7, Biolegend), and RB705-anti-PD-1 (clone EH12.1, BD Biosciences) for 30 minutes at 4°C before fixation and permeabilization with eBiosciences Foxp3/Transcription Factor Staining kit (Thermo) as per manufacturer’s protocol, intracellular staining for FITC-anti-granzyme B (clone GB11, Biolegend), and intranuclear staining for PE-anti-TCF1 (clone C63D9, Cell Signaling Technology). Cells were analyzed by flow cytometry. Samples with viability below 50% or fewer than 50 live HIV-specific pHLA-tetramer^+^ CD8^+^ T cells were excluded from downstream analyses.

### Expanded antigen-specific elimination assay

Recall cytotoxicity of HIV-1 epitope-specific memory CD8^+^ T cell responses was measured using the expanded antigen-specific elimination assay (EASEA) as per our published protocol^33^, for all proliferative responses (>5% CFSE-low) and a representative sampling of nonproliferative responses that were immunodominant based on IFN-γ elispot. Rather than per-cell cytotoxicity, this assay measures per-response endpoint cytotoxicity following *in vitro* recall. Briefly, PBMCs were rested overnight in R10 then incubated with 100 ng/ml individual HLA-optimal HIV-1 peptide for six days to expand antigen-specific effector cells. Target CD4^+^ T cells were isolated from PBMC by negative magnetic separation (StemCell Technologies), activated in 24-well non-treated polystyrene plates (Corning) pre-coated with 2 mg/ml anti-CD3 (clone OKT3, Biolegend) at 1-2 million cells/ml in R10 with 2 mg/ml anti-CD28 (clone CD28.2, Biolegend) and 50 U/ml IL-2 (Peprotech) at 37 C overnight, then expanded in treated 24-well plates (Corning) at 2 million cells/ml in R10 with 50 U/mL IL-2 at 37 C for five days. 50% of target cells were pulsed for 30 minutes at 37 C with 1 µM peptide and labeled with CellTrace Far Red dye (Thermo Fisher) and mixed with unpulsed target cells 1:1, then labeled with CellTrace Violet dye (Thermo Fisher). After six days of expansion, CFSE-labeled effector CD8+ T cells were isolated from pooled mononuclear cells by negative magnetic separation (StemCell Technologies) and co-cultured with target cells at effector:target (E:T) ratios of 0:1, 1:1, 2:1, 4:1, and 8:1 with 50,000 target cells/well in a treated 96-well polystyrene plate (Corning) for 4 hours. Effector-only populations were stained with APC-conjugated pHLA tetramers and all samples were stained with BV605-conjugated anti-CD3 (clone SK7, Biolegend), BUV395-conjugated anti-CD8 (clone RPA-T8, BD Biosciences), BV711-conjugated anti-CD4 (clone RPA-T4, Biolegend) and Live/Dead Near-IR (Thermo Fisher) then analyzed by flow cytometry. Results were gated as described previously and percent elimination and AUC were calculated as described previously^6, 33^. Responses above 100 AUC, equivalent to the first-quartile recall cytotoxicity of immunodominant HIV-specific CD8^+^ T cell responses from spontaneous controllers, were considered to have high recall cytotoxicity.

### Infected cell elimination assay

The assay described above was adapted to measure elimination of infected cells by proliferative HIV-1 Gag-specific responses from HEAL participants and spontaneous controllers via the following protocol modifications. PBMCs were labeled with CellTrace Violet dye (Thermo Fisher) instead of CFSE to avoid interference with GFP. CD4^+^ T cells were infected with VSV-G-pseudotyped HIV_NL4-3ΔEnv-EGFP_ (NIH AIDS Reagent Program, Division of AIDS, NIAID, NIH, Cat#11100^65^) for the final 3 days of culture. Prior to coculture with autologous CD8^+^ T cells, productively infected cells with downmodulated CD4 were enriched by magnetic CD4 depletion (EasySep CD4 Positive Selection Kit, StemCell Technologies) and mixed with autologous uninfected CD4 T cells to a final infection rate ∼30%, then labeled with CellTrace FarRed. Cocultures were incubated for 16 hours to enable sufficient sensitivity. Cells from PWOH were stimulated with an A*02-restricted CMV NLVPMVATV and were included as negative controls to estimate assay sensitivity.

### HIV-1 reservoir measurements

Genomic DNA and total RNA were isolated from approximately 5x10^6^ peripheral blood mononuclear cells (PBMC) or total CD4^+^ T cells (AllPrep DNA/RNA kit, Qiagen). Total HIV-1 DNA and unspliced cell-associated RNA (usRNA) levels were quantified in triplicate by real-time PCR as previously described, modified to include Taqman Universal Mastermix (DNA) or Taqman Fast Virus 1-Step Mastermix (RNA)^66^. HIV-1 quantifications used forward primer 5’-TACTGACGCTCTCGCACC-3’, reverse primer 5’-TCTCGACGCAGGACTCG-3’, and probe 5’ FAM-CTCTCTCCTTCTAGCCTC-MGB 3’ (ThermoFisher Scientific). DNA cycling conditions in a total reaction volume of 25µL were 95°C for 15 min followed by 40 cycles of 95°C for 15 sec and 60°C for 1 min. RNA cycling conditions in a total reaction volume of 20µL were 55°C for 15 min, 95°C for 20 sec, followed by 40 cycles of 95°C for 3 sec and 60°C for 30 sec. Approximately 500ng of genomic DNA were assessed per well. For polyadenylated HIV-1 mRNA quantifications, 10uL (approximately 1 million cell equivalents) of RNA were run in triplicate using primers and probe targeting the nef to poly-A region of HIV-1 mRNA as previously described^67^. The limits of quantification for total HIV-1 DNA and unspliced caRNA were 1 and 3 copies per reaction, respectively. Limits of detection were calculated per timepoint and participant sample, considering the average number of copies detected across triplicate measurements. Cell input numbers were quantified by human genome equivalents of CCR5 DNA using forward primer 5’-ATGATTCCTGGGAGAGACGC-3’, reverse primer 5’-AGCCAGGACGGTCACCTT-3’, and probe 5’ FAM-CTCTCTCCTTCTAGCCTC-MGB 3’ (ThermoFisher Scientific) as described^66^. To calculate copies of HIV-1 caRNA per million cells, copy numbers were averaged and cell counts normalized to a CCR5 DNA reference. Our HIV-1 unspliced and poly(A) caRNA amplifications are both one-step qPCR assays that use an HIV-specific primer to synthesize cDNA. For poly(A) HIV-1 mRNA quantifications, copies per million cell equivalents were calculated by initial cell count prior to reactivation. Both HIV-1 usRNA and polyA assays used Taqman Fast Virus 1-Step Mastermix in a total reaction volume of 20μL and identical cycling conditions.

HIV-1 DNA from isolated PBMC DNA was measured using the cross-subtype intact proviral DNA assay (CS-IPDA) as described^68, 69^, with minor modifications. Two multiplexed droplet digital PCR assays (Bio-Rad QX200) were performed per sample targeting conserved HIV-1 sequences and, to normalize data, a cell reference gene. Master mixes were prepared using 2X ddPCR Supermix for probes. The three primer/probe sets of cross subtype primers used in this study, which have been shown to bind to target regions of Subtype B, A, C, D^68^, CS-IPDA 5’pol (Forward: 5’-WCCYTTARYTTCCCTCARATCACTCT-3’; Reverse: 5’-TACTGTATCATCTGCTCCTGTRTCTAAKAGAGCYTC-3’; Probe: 5’-TTGGCARCGACC-3’ FAM, BHQ); CS-IPDA LTR/Gag (Forward: 5’-GACTAGCGGAGGCTAGAAGGAGAGA-3’; Reverse: 5’-CTAATTTTCCSCCDCTTAATAYTGACG-3’; Probe: 5’-ATGGGTGCGAGA-3’ HEX, BHQ); and CS-IPDA Env (Forward: 5’-TVTTCMTTGGGTTCTTRGGAGCAGCAGG-3’; Reverse: 5’-GCACTATRCCAGACAATAVYTGTCTGGCCTGTACC-3’; Probe: 5’-AGCACKATGGG-3’; HEX, BHQ). To estimate the number of cells per reaction and correct for sheared DNA, separate reactions targeting two regions of the *RPP30* gene, RPP30-1 (Forward: 5’-GATTTGGACCTGCGAGCG-3’; Reverse: 5’-GCGGCTGTCTCCACAAGT-3’; Probe: 5’-CTGACCTGAAGGCTCT-3’, HEX, MGB) and RPP30-2 (Forward: 5’-GACACAATGTTTGGTACATGGTTAA-3’; Reverse: 5’-CTTTGCTTTGTATGTTGGCAGAAA-3’; Probe: 5’-CCATCTCACCAATCATTCTCCTTCCTTC-3’, FAM). Droplets were generated and subjected to PCR cycle: 95°C for 10 minutes, followed by 60 cycles of 94°C for 30 seconds, 59°C for 1 minute, and ending with 98°C for 10 minutes before holding at 4°C. To increase the assay sensitivity, we run CS-IPDA in multiple wells with the DNA from minimum of cell 500,000 cells. Data from replicate wells were summed and analyzed using QuantaSoft Data Analysis Software. Droplets containing a triple positive signal (*pol*+/*gag*+/*env*+) were considered intact, and all other droplets were considered defective with deletions. Relative DNA shearing was calculated using the ratio of single positive RPP30 droplets compared to the double positive population and was used to estimate the number of intact proviruses. The number of diploid cells in each reaction was determined by dividing the copies of RPP30 in half and correcting for the dilution factor for the HIV-1 reaction. Intact, defective, and total HIV-1 DNA were normalized to the number of cells and reported as copies/million PBMC. 11 participants with detectable plasma viral loads within one year of sampling were excluded from reservoir analyses.

### Whole-PBMC single-cell multiomics

Cryopreserved PBMCs from longitudinal samples collected during ATI from participant H047 were thawed and rested overnight at 37°C in media that contained raltegravir 2 µM and tenofovir 2 µM (Selleckchem). Cells were then washed with Cell Staining Buffer (BioLegend) and incubated with Fc receptor blocking solution (Human TruStain FcX, Biolegend) for 10 minutes at 4°C. Cells from individual time points were barcoded by unique hashtag oligonucleotide (HTO) antibodies (Biolegend) for 20–30 min at 4 °C and incubated with Sytox Blue Dead Cell Stain (Invitrogen) at room temperature for 15 min before live cell sorting (SONY SH800, Ragon Institute). Sorted live cells were subsequently incubated with TotalSeq-C Human Universal Cocktail V2.0 (BioLegend) for 30 min at 4 °C and washed three times with Cell Staining Buffer. Stained and barcoded cells were immediately loaded onto Chromium X instrument (10X Genomics) for library construction for T and B cell receptor variable-diversity-joining (V(D)J) diversity analysis, 5’ gene expression and cell surface protein profiling.

Sequencing data for gene expression, antibody capture, and BCR/TCR V(D)J were processed using the Cell Ranger multi pipeline (10X Genomics, v 9.0.0) with the GRCh38 human reference genome, TotalSeq-C feature reference sequences, and human V(D)J reference sequences, respectively. Demultiplexed data from each sample were imported into R (v 4.4.0) and combined for joint analysis using Seurat (v 5.2.1). A total of 37,739 PBMC before filtering and 37,462 PBMC after filtering were obtained for subsequent analyses. RNA filtering criteria were: 1) number of UMIs (nCount_RNA) > 1500, 2) number of detected genes (nFeature_RNA) > 1000, 3) mitochondrial percentages (percent_mito) < 20, and 4) novelty (novelty) > 0.8. ADT filtering criteria were number of UMIs (nCount_ADT) > 400 and number of detected proteins (nFeature_ADT) > 100.

RNA data were normalized using SCTransform^70^, and TCR V(D)J genes were excluded from the set of highly variable features prior to principal component analysis (PCA). A uniform manifold approximation and projection (UMAP) embedding was generated using the top 30 principal components. The antibody-derived tag (ADT) counts were normalized by centered log ratio (CLR) transformation, followed by PCA and UMAP. RNA and ADT modalities were then integrated using a weighted nearest neighbor (WNN) algorithm^71^, and UMAP and clustering was computed on the WNN graph.

TCR and BCR data were imported into R and incorporated into the Seurat object using scRepertoire^72^. TCR clonotypes were defined by the CDR3 amino acid sequence of the beta chain, and BCR clonotypes were defined using scRepertoire’s strict definition, which is based on the normalized Levenshtein edit distance of CDR3 nucleotide sequences and V-gene usage. Clonal proportions were calculated as the relative abundance of each clonotype within the total immune repertoire of the sample.

Cell types were assigned using a reference-mapping approach with the Azimuth PBMC reference. CD8^⁺^ T cells, CD4^⁺^ T cells, B cells, and Natural Killer (NK) cells were subsequently subsetted and reanalyzed. Differentially expressed genes (DEGs) were identified using the FindMarkers function in Seurat.

### HIV-specific CD8^+^ T cell single-cell multiomics

Cryopreserved H047 PBMC were thawed and rested overnight before negative-selection magnetic CD8^+^ T cell isolation (StemCell Technologies), then stained with APC-conjugated HLA-B*44 AEQASQEVKNW tetramer, Total-Seq C Human Universal Cocktail (Biolegend), and six unique Total-Seq C hashing antibodies (Biolegend) corresponding to biweekly sample time points during the treatment interruption (TI) following reinitiation of ART (RA). 40,490 sorted cells were encapsulated in GEM-wells using a Chromium controller (10X Genomics). Gene expression (GEX), surface protein expression (antibody-derived tags, ADT), and TCR (VDJ) libraries were generated using the 10X Chromium NextGEM Single Cell 5’ v2 Dual Index kit with feature barcoding technology (10X Genomics) following the manufacturer’s instructions. Libraries were pooled at a 5:1:1 GEX:ADT:VDJ ratio and sequenced on a NextSeq 2000 instrument with a 100-cycle P3 kit (Illumina).

Base-calling was performed using bcl2fastq and initial data-processing was performed via the Cell Ranger multi pipeline (10X Genomics, v 7.0.0) using refdata-gex-GRCh38-2020-A as a transcriptome reference and refdata-cellranger-vdj-GRCh38-alts-ensembl-5.0.0 as a VDJ reference. Gene expression (GEX), antibody capture (ADT), and TCR (VDJ) libraries were specified in the multi analysis config file. Surface protein barcodes and hashtag barcodes corresponding to time points were designated as “Antibody Capture” in the feature-reference file. After processing by Cell Ranger, the count matrix in sample_filtered_feature_bc_matrix was analyzed using the Seurat R package. Hash-tag level timepoint sample demultiplexing was performed using the HTOdemux() function of Seurat, and cells were removed for which HTO_classification.global was not “Singlet”, hence removing cells with multiple or no hashtags. 9,947 AW11-specific cells were recovered, of which 7,500 passed filtering, to include 493 in sample TI0, 1872 in TI2, 866 in TI4, 916 in TI6, 1625 in AR2, and 1728 in AR4. The GEX library yielded 45,823 median reads, 1,787 median genes, and 3,778 median UMIs per cell. The ADT library yielded 1,375 median ADT UMIs per cell. The VDJ library yielded 6,663 cells with productive V-J spanning pairs, and 3 and 17 median TRA and TRB UMIs per cell, respectively. To avoid clustering driven by clonotype-specific TCR gene expression, gene features for which the symbols matched the regular expression “^TR[ABDG][VJC]” were removed from the data set prior to clustering^73^. Using the Seurat function FindVariableFeatures(), 4,000 variable genes were selected for dimensionality reduction and differential expression analysis. Counts were log normalized, scaled and centered prior to dimensionality reduction and clustering. Clustering was performed using SNN clustering via Seurat’s Seurat::FindNeighbors FindClusters() function with the argument resolution = 0.35. TCR clonotypes were assigned on the basis of raw_clonotype_id from the vdj_tfiltered_contig_annotations.csv file generated by Cell Ranger. Differential expression was performed using Seurat’s FindMarkers() function. Pathway analysis was performed using the tmodCERNOtest() from the tmod R package version 0.46.2^74^ using a subset of MSigDB version v7.5.1^75^ that included the hallmark (H), gene ontology, reactome, and KEGG gene sets. Gene set network analysis was performed using the GSNA R package, version 0.1.4.9, as previously described^6, 76^.

### Flow cytometry

All flow cytometry data was acquired at the Ragon Institute flow cytometry core facility using BD Symphony, Fortessa, and LSR-II cytometers. Fluorescence spillover compensation was applied using FACSDiva software (BD Biosciences) after collection of single-stained cell- or bead-based compensation controls (Thermo Fisher). Data was analyzed using FlowJo v10.10. Gating thresholds were established using appropriate negative controls, including fluorescence-minus-one and biological negative controls. Gates were applied consistently across all intra- and inter-sample comparisons.

### Statistical analyses

Statistical analyses were performed using GraphPad Prism and R (vers.4.4.0). Assuming a general non-Normal distribution of the data, non-parametric tests based on ranks were used to compare continuous measurements across groups (Wilcoxon rank-sum tests), as well as categorical variables (Fisher’s exact tests), where needed. For the same reason, Spearman’s correlation coefficients (with corresponding tests) were preferred to Pearson’s, to verify monotonicity between measurements.

## Data Availability

All data produced in the present study are included in the article's supplementary information or available upon reasonable request to the authors.

## Resource availability

Single-cell multiomics are available via NCBI Gene Expression Omnibus (GEO) accession numbers GSE294440 and pending. The remaining data are included in the manuscript and supplementary materials.

## ACKNOWLEDGMENTS

The authors are grateful to the study participants; to the American Red Cross HLA genotyping laboratory; and to Alicja Piechocka-Trocha, Mary Carrington, Ashok Khatri, Michael Waring, Karen Power, Fernando Senjobe, Nishant Singh, Julia Hitschfel, and Elif Lakan for technical advice and assistance. This work was supported by funding from United States National Institutes of Health (NIH) R01 AI149704, R01 AI157854, R01 AI172843, DP2 AI184606, P01 AI169768, P30 AI042853, P30 AI060354, T32 AI007387, L30 AI126472, and Howard Hughes Medical Institute. This project was further supported by Clinical Translational Science Award 1UL1TR002541 to Harvard University and Brigham and Women’s Hospital from the National Center for Research Resources. The content is solely the responsibility of the authors and does not necessarily represent the official views of the National Center for Research Resources or the National Institutes of Health. Funders had no role in study design, data collection and analysis, decision to publish or preparation of the manuscript.

## Author contributions

Conceptualization: DRC, AT

Methodology: DRC, LCW, KLC, MS, SZ, AT

Investigation: DRC, MJO, SZ, TJL, RR, XH, JYC, BC, UA, ZJR, HW, CC, XG, JMD

Data curation: HCJ, JMU, ZZ, NS, JB, DRC, AT

Formal analysis: EM, JMU, DRC, ZZ, NS, JB

Resources: SHS, BDW, AT

Writing – original draft: DRC

Writing – review and editing: All authors

Funding acquisition: DRC, BDW, MS, AT

Supervision: DRC, MS, SHS, BDW, AT

## LIST OF SUPPMENTARY MATERIALS

Extended Data Figs. 1 to 9

Extended Data Tables 1 to 2

Supplementary Data 1 to 3

