## Extended Data for "CD8^+^ T cell recall cytotoxicity during antiretroviral therapy is associated with limited HIV-1 reservoir size and activity"

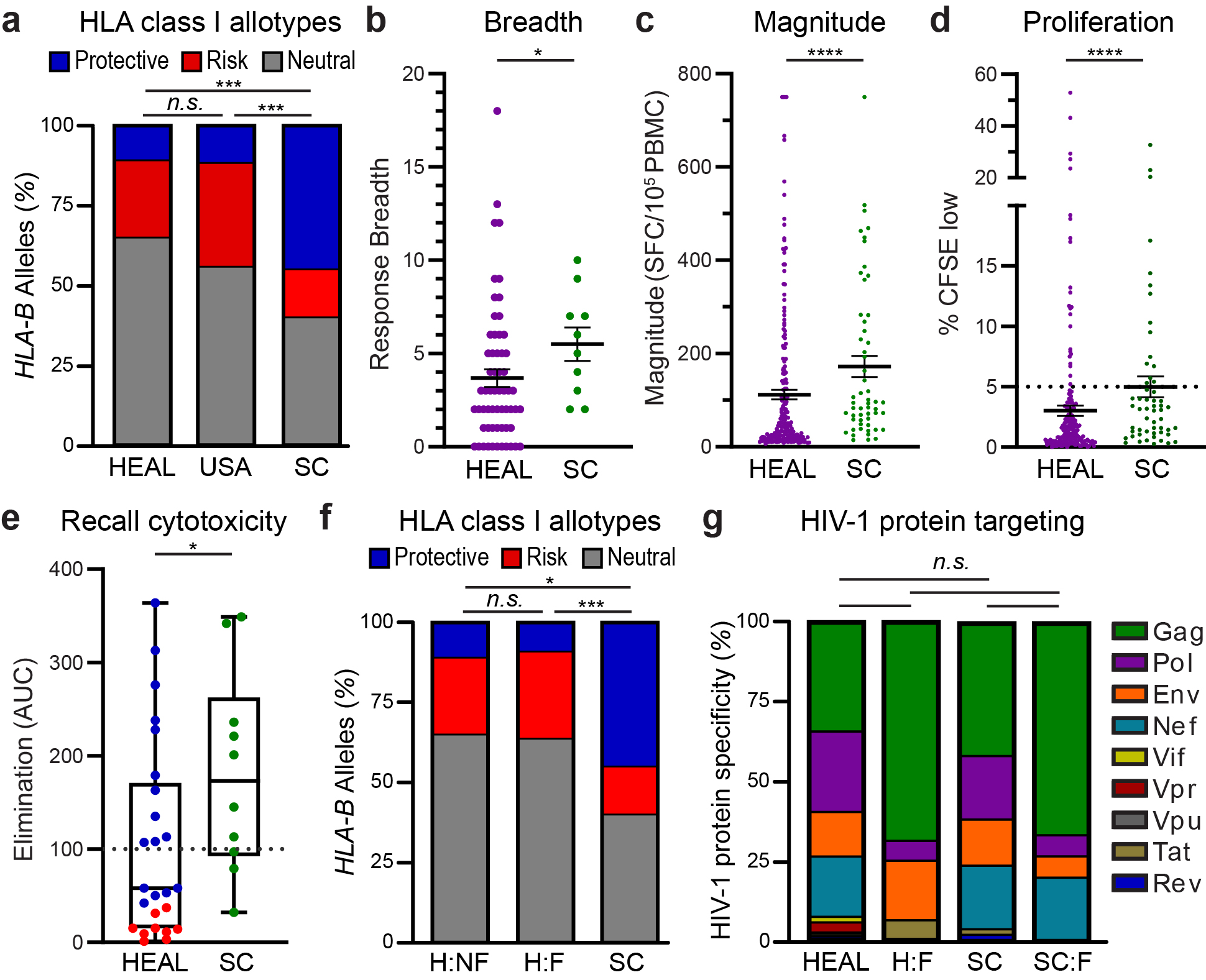

**Extended Data Fig. 1: Comparisons of HEAL participants to spontaneous HIV-1 controllers. (a)** *HLA-B* allelic frequencies among 60 HEAL participants compared to the national United States population (*n.s.*, not significant; *p*>0.05) or spontaneous HIV-1 controllers (SC; ***, p<0.001, χ^2^ tests) controlling for race and ethnicity, grouped by prior GWAS association with HIV-1 viral loads during untreated natural infection: protective (*HLA-B*14*, *B*27*, *B*52*, *B*57*), risk (*HLA-B*07*, *B*08*, *B*35*), and neutral (all other *HLA-B* alleles). **(b-d)** Number of unique epitope-specific responses (b) and magnitude (c; SFC, spot-forming counts) measured by IFN-γ elispot, proliferation measured by CFSE dilution (d) in HEAL (*n=*221 responses from 60 participants) and spontaneous HIV-1 controllers (SC, *n*=55 responses from 10 participants). Solid lines represent means +/-SEM. Dotted line represents threshold set to mean of SC group (5%). * *p*<0.05, *****p*<0.0001, Wilcoxon signed-rank test. **(e)** Recall cytotoxicity (AUC, area under curve) of HIV-specific CD8^+^ T cells from HEAL participants with high (blue, >5% CFSE-low, *n=*16 responses from 14 participants) or low (red, <5% CFSE, 9 responses from 8 participants) proliferative capacity or immunodominant responses from spontaneous HIV-1 controllers (SC, *n*=10 responses from 10 participants). Box and whiskers represent median, IQR, and range. Dotted line represents threshold set to first quartile of SC group (100 AUC). **p*<0.05, Wilcoxon signed-rank test. **(f)** *HLA-B* allelic frequencies, as in a, for subsets of HEAL (H:) participants with nonfunctional (NF; *n=*50) and functional (F, *n*=10) recall cytotoxicity, defined as in Fig. 2, and for spontaneous HIV-1 controllers (SC, *n*=10). *n.s.* *p*>0.05, **p*<0.05, ****p*<0.001, χ^2^ tests. **(g)** HIV-1 proteins for which CD8^+^ T cell responses are specific among total cohorts and subsets with functional CD8^+^ T cell responses (:F) among HEAL participants (*n=*221 responses from 60 participants) or spontaneous HIV-1 controllers (SC; *n*=55 responses from 10 participants). All *n.s.* *p*>0.05, χ^2^ tests.

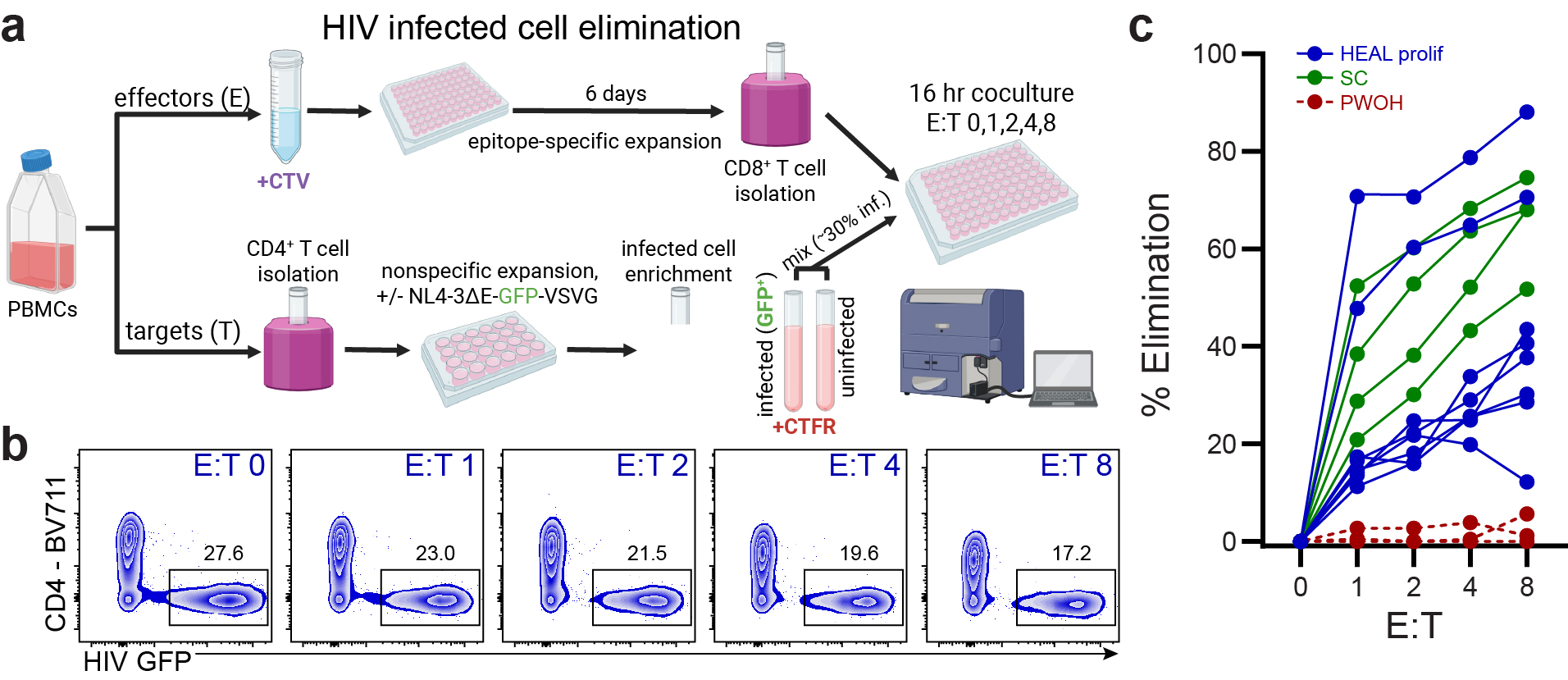

**Extended Data Fig. 2: HIV-infected cell elimination. (a)** Experimental design for measurement of infected CD4^+^ T cell elimination by autologous peptide-expanded CD8^+^ T cells, as described in Methods. CTV, CellTrace Violet; CTFR, CellTrace Far Red. **(b)** Elimination of HIV GFP^+^ cells at increasing effector:target (E:T) ratios by a representative HLA-B*14-restricted Gag DA9-specific CD8^+^ T cell response from participant H086. **(c)** Percent elimination of infected target cells for proliferative Gag-specific CD8^+^ T cell responses from HEAL (blue, *n*=8) and spontaneous controller (SC, green, *n*=4) participants, and negative controls from people without HIV (PWOH, *n*=3).

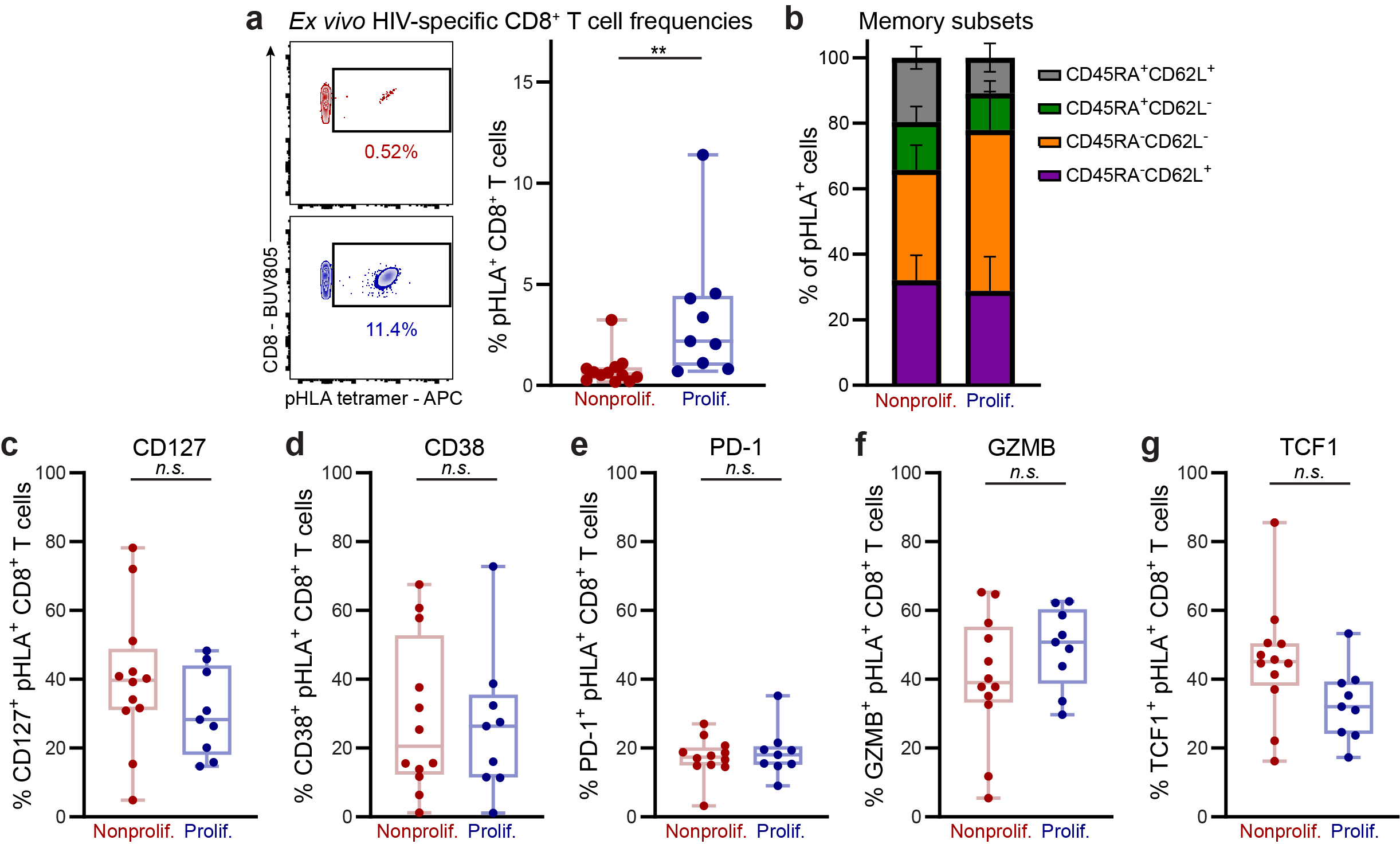

**Extended Data Fig. 3: Flow cytometric phenotyping of nonproliferative and proliferative HIV-specific CD8^+^ T cells. (a)** Left: Examples of *ex vivo* tetramer staining with HIV-1 peptide-HLA (pHLA) tetramers, including for HLA-C*08 Gag TPQDLNTML in HEAL participants H060 (red; poor proliferative capacity) and H017 (blue, high proliferative capacity). Right: Summary of *ex vivo* immunodominant HIV-1 pHLA tetramer staining of nonproliferative (red, <5% CFSE low, *n*=12 responses from 10 participants) and proliferative (blue, ≥5% CFSE low, *n*=9 responses from 9 participants) CD8^+^ T cells. ***p*=0.0013, Wilcoxon rank-sum test. **(b)** Summary of memory subset frequencies among HIV-1 pHLA tetramer^+^ CD8^+^ T cells as defined by CD45RA and CD62L staining among responses from participants with low (<5%) or high (≥5%) proliferative capacity, as measured in Fig. 1D. Bars represent means +/- SEM. **(c-g)** Percent of HIV-1 pHLA tetramer^+^ CD8^+^ T cells expressing CD127 (c), CD38 (d), PD-1 (e), granzyme B (f), and TCF1 (g) among responses from participants with low (<5%) or high (≥5%) proliferative capacity, as measured in Fig. 1d. *p* values calculated by Wilcoxon rank-sum tests.

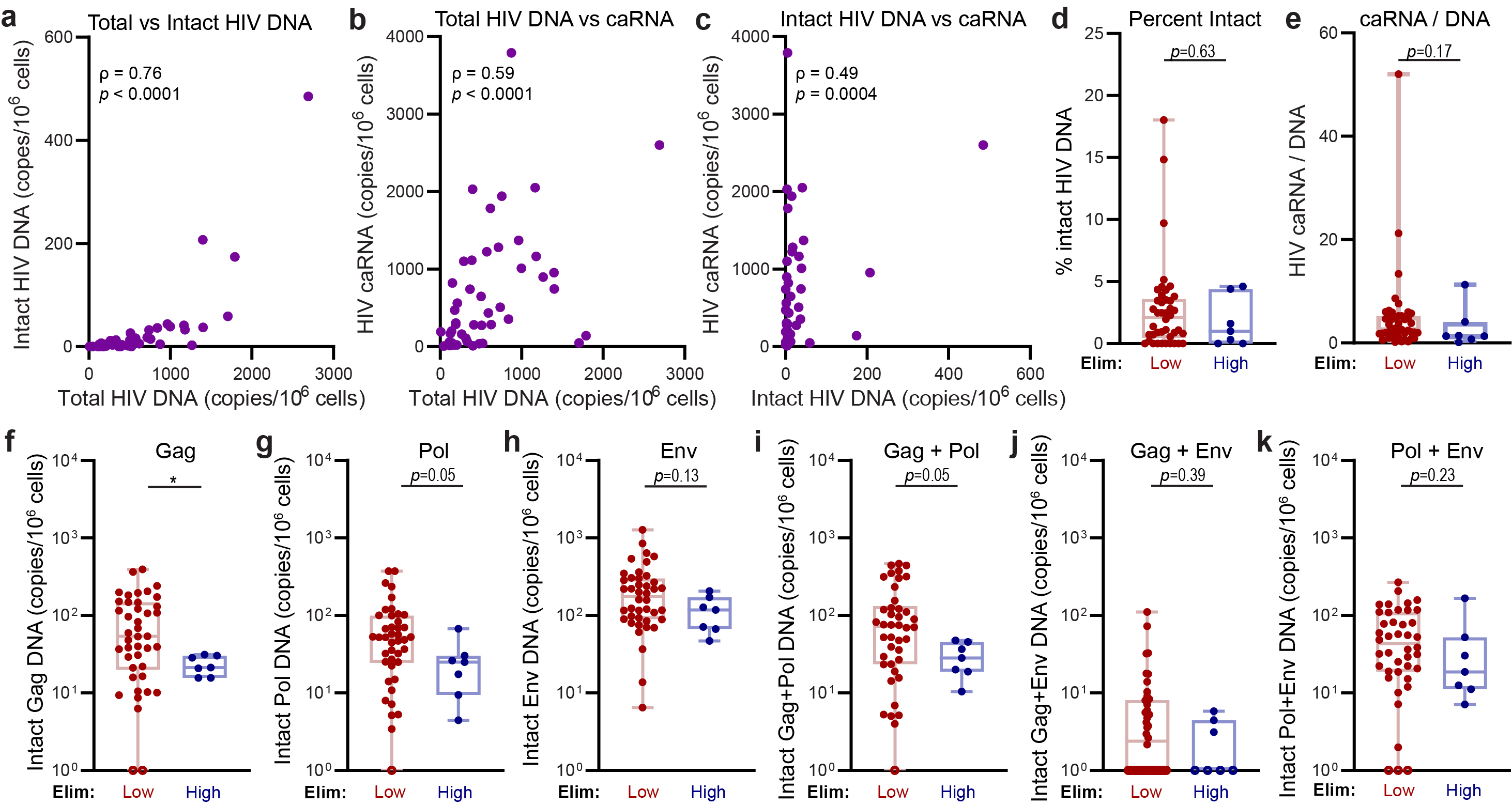

**Extended Data Fig. 4: Extended HIV reservoir analyses. (a-c)** Spearman correlations (ρ) with corresponding correlation tests of total versus intact HIV-1 DNA (a), total HIV-1 DNA vs caRNA (b), and intact HIV-1 DNA vs caRNA (c), *n*=49 HEAL participants. **(d-k)** Percent of intact relative to total HIV-1 proviral DNA (d), ratio of HIV-1 caRNA to DNA (e), intact HIV-1 Gag DNA (f), intact Pol DNA (g), intact Env DNA (h), intact Gag+Pol DNA (i), intact Gag+Env DNA (j), Intact Pol+Env DNA (k) from participants with HIV-specific CD8^+^ T cell responses of low (max. elim AUC <100, *n*=42) or high (max. elim AUC >100, *n*=7) cytolytic capacity. Open circles at 10^0^ represent values below the assay detection limit. **p* < 0.05; *p* values calculated by Wilcoxon rank-sum tests.

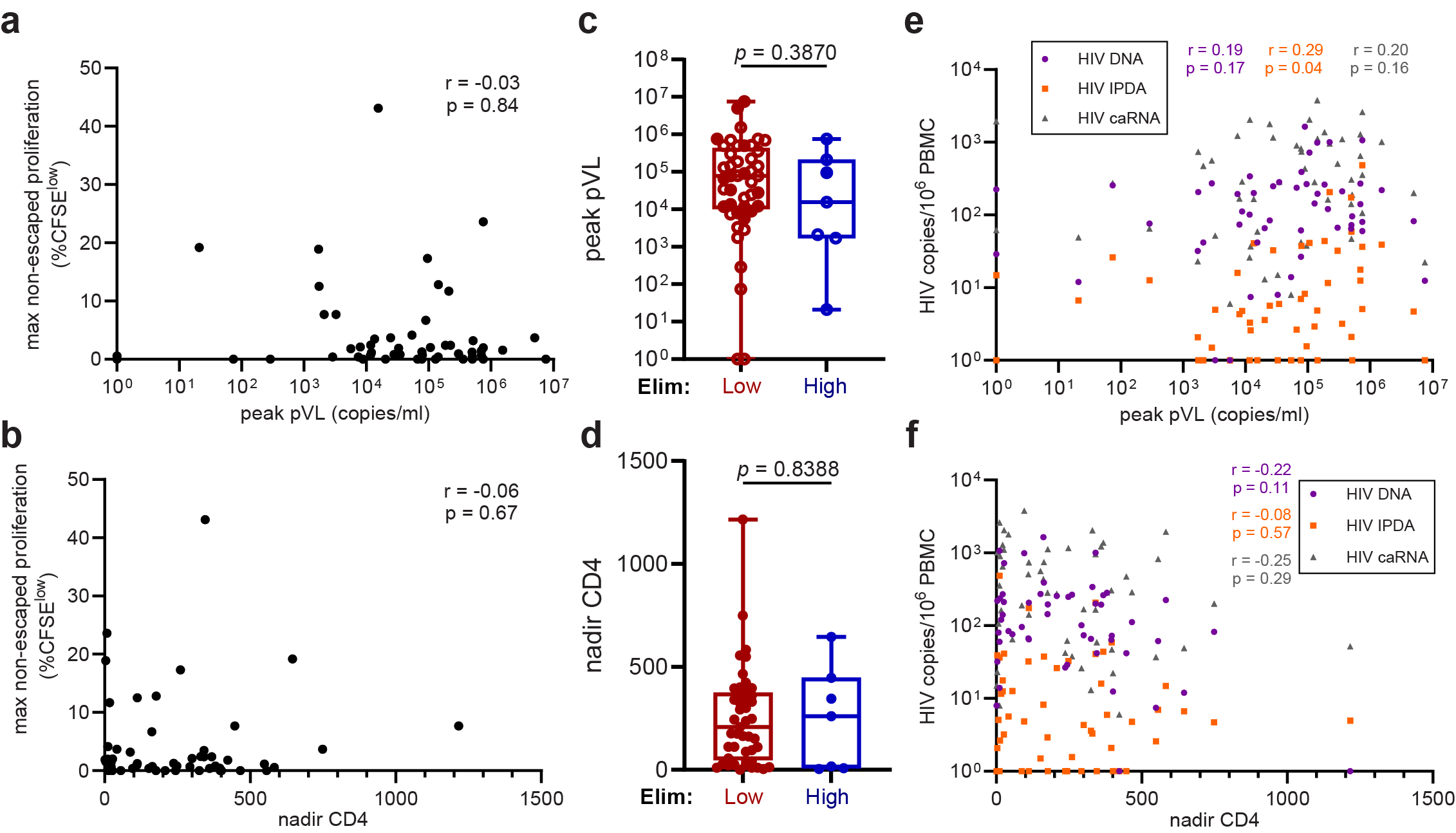

**Extended Data Fig. 5: Lack of associations between clinical metrics of disease progression and CD8^+^ T cell functionality or reservoir size.** **(a-b)** Spearman correlations of peak plasma viral load (pVL, a) or nadir CD4 (b) and maximum non-escaped HIV epitope-specific CD8^+^ T cell proliferation among HEAL participants. **(c-d)** Peak pVL (c) and nadir CD4 (d) among HEAL participants stratified by low (AUC≤100, *n=*42) or high (AUC>100, n*=7*) recall cytotoxicity. Open circles represent pVL values obtained post-ART initiation. **(e-f)** Spearman correlations of peak pVL (e) or nadir CD4 (f) and total HIV-1 DNA, intact HIV-1 proviral DNA (IPDA), or cell-associated HIV-1 RNA (caRNA) among HEAL participants. *n*=49 participants.

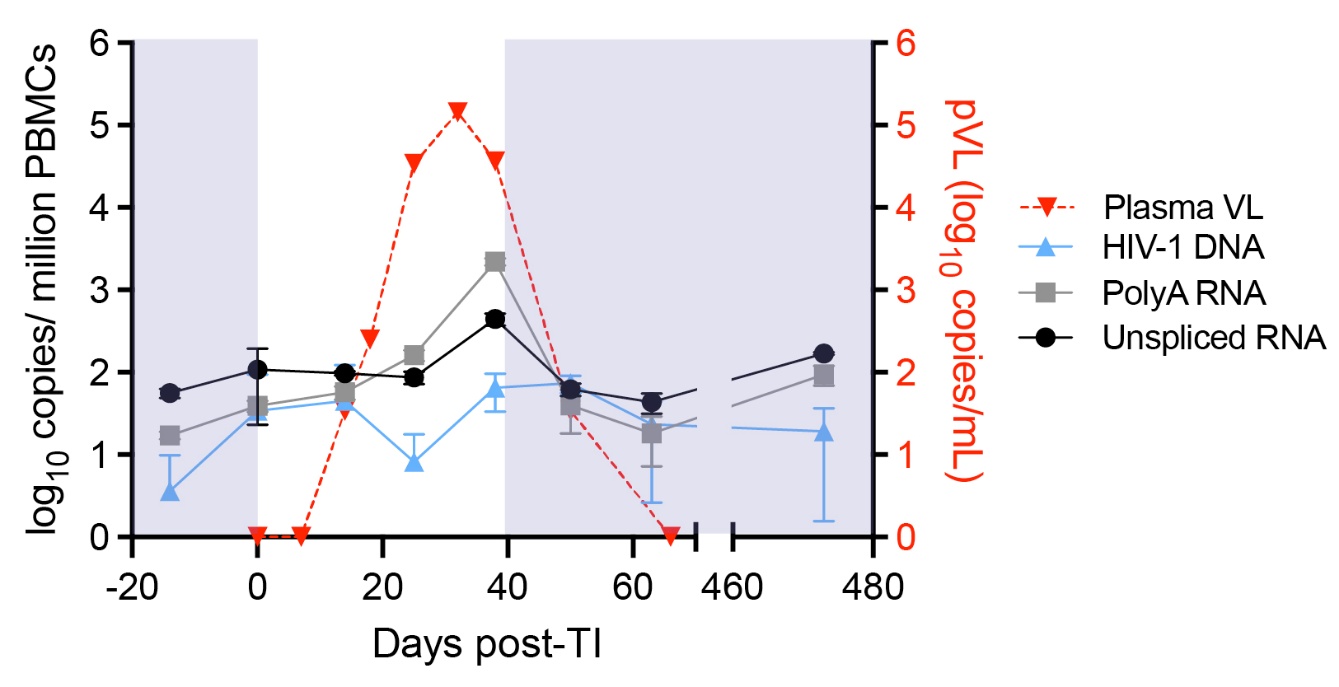

**Extended Data Fig. 6: HIV-1 persistence markers during ATI.** Longitudinal plasma viral load (VL), total HIV-1 DNA, polyadenylated (PolyA) and unspliced HIV-1 RNA quantitation during analytical treatment interruption (ATI) in participant H047. Triplicate technical replicates were performed and averaged; error bars represent standard deviations. Shaded boxes represent times when the participant was on ART. Please note that the study visit where peak HIV-1 plasma viremia was observed did not, per protocol, include a PBMC collection.

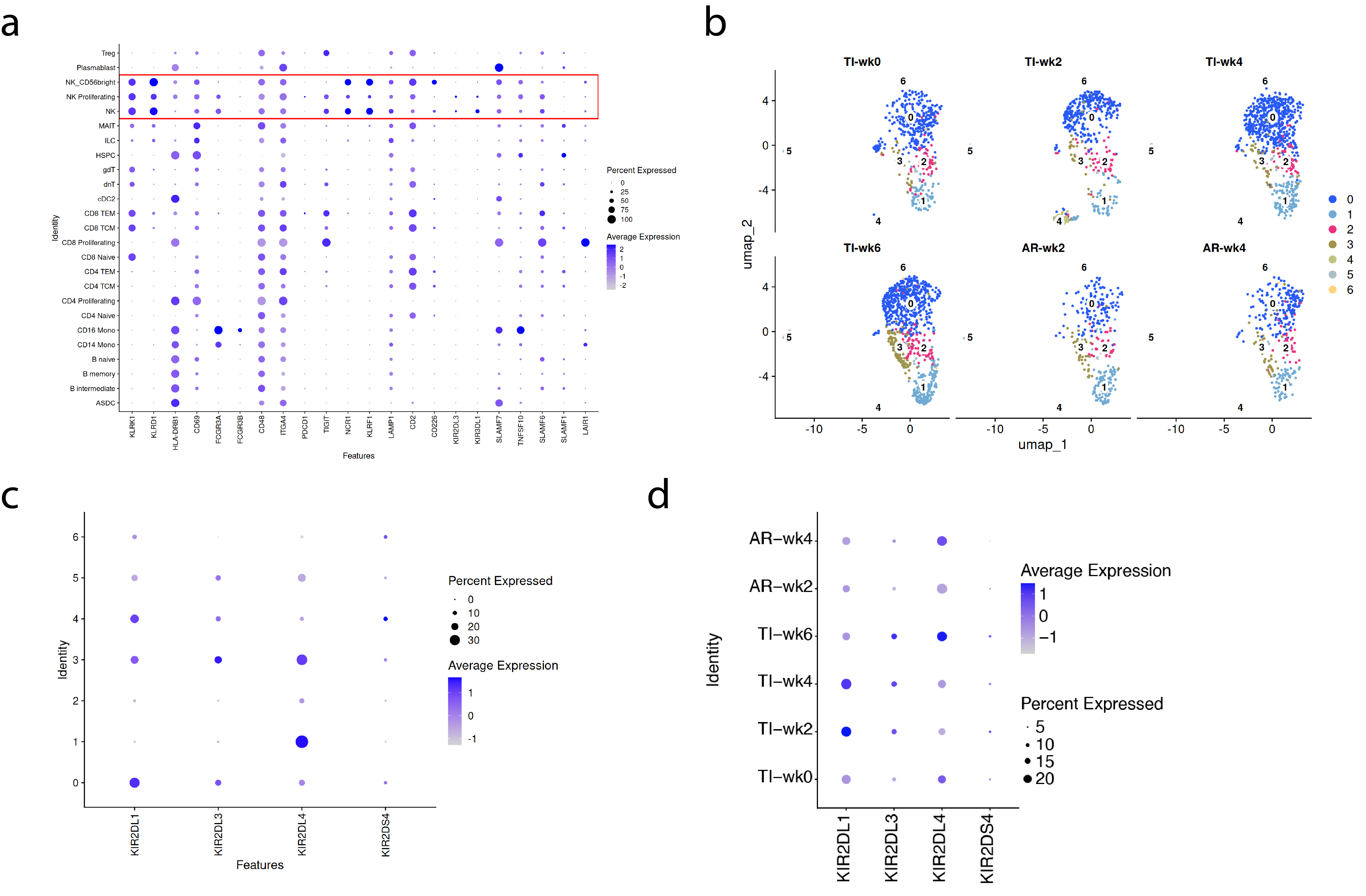

**Extended Data Fig. 7: NK cell expansion during ATI.** **(a)** Dot plot that displays the percent expression and average expression levels of NK cell phenotyping marker transcripts in H047 PBMC subpopulations, as identified in the WNN UMAP of Fig. 3b. The red box highlights the three identified NK cell types. **(b)** Longitudinal NK cell subclustering analysis at resolution 0.2. **(c-d)** Dot plots of KIR family genes contained in the dataset, as a function of NK cell subcluster (c) or ATI sample timepoint (d).

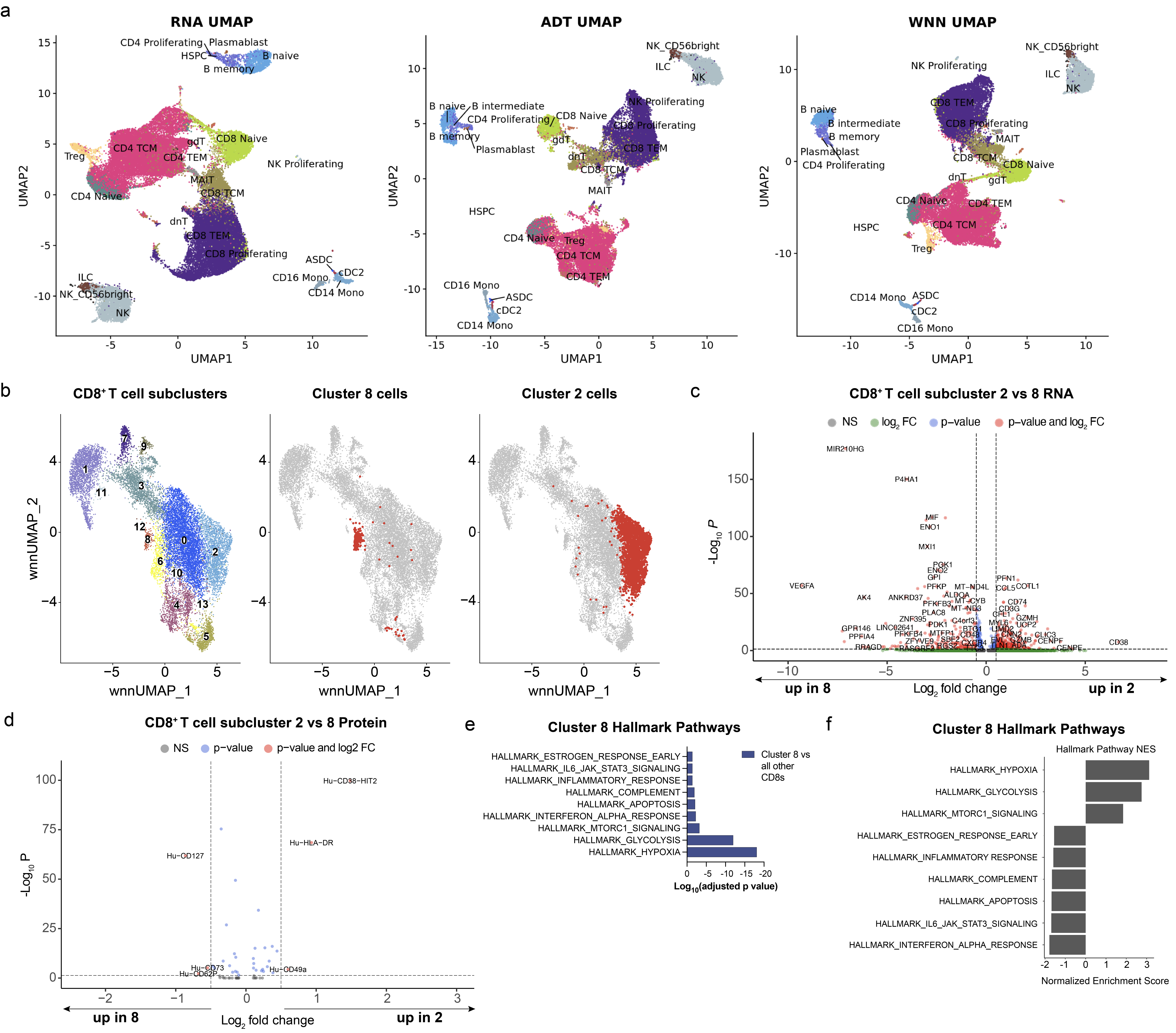

**Extended Data Fig. 8: CD8^+^ T cell characterization during ATI. (a)** RNA, ADT, and WNN UMAP dimensionality reductions for 5’ CITE-seq performed with longitudinal PBMC samples collected before, during, and after an antiretroviral treatment interruption in participant H047. Note the WNN panel shown here is identical to the WNN UMAP displayed in Fig. 3b. **(b)** CD8^+^ T cell subclustering analysis of total CD8^+^ T cells from all ATI samples as displayed in Fig. 3D, highlighting clusters 8 and 2 that changed longitudinally. Note that cluster 8 was observed only in sample TI2. **(c-d)** Volcano plots summarizing differential gene (c) and surface protein (d) expression between CD8^+^ T cell subclusters 2 and 8. **(e)** Gene set enrichment analysis (GSEA) of the top statistically significantly different Hallmark gene sets between cluster 8 and all other CD8+ T cells, **(f)** Magnitude of the Hallmark gene sets’ differential expression, displayed as normalized enrichment scores. NES >0 indicates gene sets enriched in upregulated genes, whereas NES<0 denotes gene sets enriched in downregulated genes.

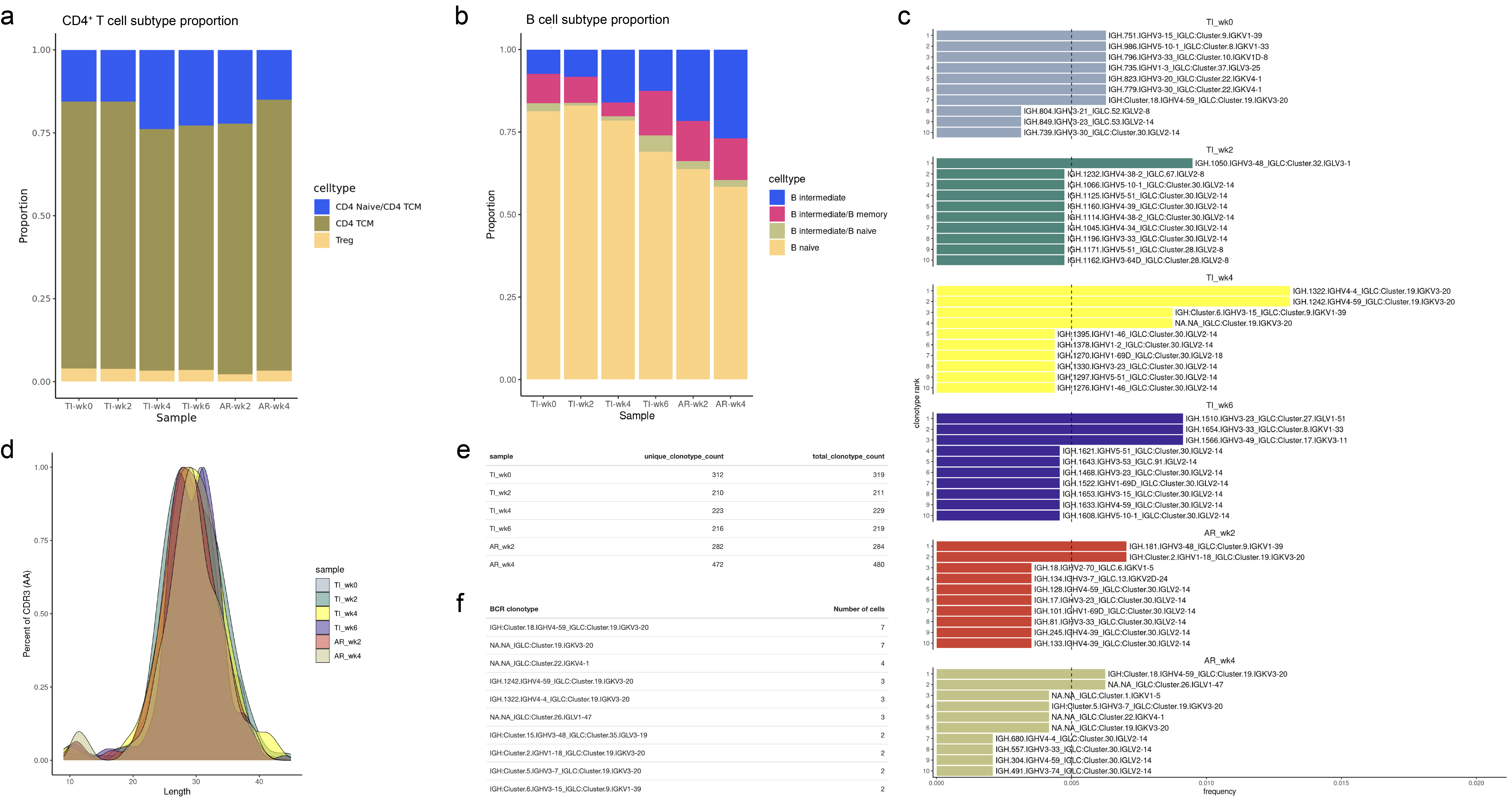

**Extended Data Fig. 9: Longitudinal CD4^+^ T cell and B cell changes during ATI.** **(a-b)** Stacked bar plots displaying the proportions of CD4^+^ T cell (a) and B cell (b) subsets over time during ATI in participant H047. **(c)** BCR clonality analysis during ATI, displaying the proportions of the top 10 clonotypes at each time point. **(d)** Density distribution plots of BCR CDR3 amino acid sequence lengths as a function of time point. **(e)** The total number of BCR clonotypes and the unique clonotype count are reported per time point, demonstrating that >99% of the BCR clonotypes we detected in the ~5,000-8,000 PBMC we sequenced per timepoint were unique. **(f)** Summary of the top 10 most abundant clonotypes and the number of B cells per clonotype, emphasizing that only B cell clonal populations of very small size were seen in peripheral blood, in contrast to our observations with clonal CD8^+^ T cell subpopulations.

|  | **AUC <100 (N=42)** | **AUC >100 (N=7)** | ***p* value** |
| --- | --- | --- | --- |
| ***Treatment with integrase inhibitor*** | | | ***0.075*** |
| **No** | 5 (11.9%) | 3 (42.9%) |  |
| **Yes** | 37 (88.1%) | 4 (57.1%) |  |
| ***Peak plasma VL during ART (copies/ml)*** | | | ***0.220*** |
| *Missing obs* | *10* | *0* |  |
| **Mean (SD)** | 248698.03 (343235.44) | 139950.00 (279806.15) |  |
| **Range** | 28.00 - 1529888.00 | 21.00 - 750001.00 |  |
| ***Absolute CD4 count at ART start (cells/mm^3^)*** | | | ***0.254*** |
| *Missing obs* | *11* | *3* |  |
| **Mean (SD)** | 338.84 (220.61) | 462.00 (323.39) |  |
| **Range** | 0.00 - 908.00 | 30.00 - 800.00 |  |
| ***Duration of undetectable viremia (months)*** | | | ***0.265*** |
| **Mean (SD)** | 105.98 (60.41) | 75.00 (34.55) |  |
| **Range** | 19.00 - 253.00 | 36.00 - 130.00 |  |
| ***Age (years)*** | | | ***0.296*** |
| **Mean (SD)** | 57.33 (7.68) | 60.71 (5.88) |  |
| **Range** | 36.00 - 70.00 | 53.00 - 69.00 |  |
| ***Duration of HIV infection (years)*** | | | ***0.352*** |
| **Mean (SD)** | 23.17 (7.48) | 26.29 (5.62) |  |
| **Range** | 6.00 - 40.00 | 18.00 - 36.00 |  |
| ***Year of diagnosis*** | | | ***0.397*** |
| **1983-2000** | 26 (61.9%) | 6 (85.7%) |  |
| **2000-2017** | 16 (38.1%) | 1 (14.3%) |  |
| ***Biological sex*** | | | ***0.398*** |
| **Female** | 14 (33.3%) | 4 (57.1%) |  |
| **Male** | 28 (66.7%) | 3 (42.9%) |  |
| ***Race*** | | | ***0.408*** |
| *Missing obs* | *1* | *0* |  |
| **Asian** | 1 (2.4%) | 0 (0.0%) |  |
| **Black** | 12 (29.3%) | 4 (57.1%) |  |
| **Other** | 7 (17.1%) | 1 (14.3%) |  |
| **White** | 21 (51.2%) | 2 (28.6%) |  |
| ***Peak VL before ART (copies/ml)*** | | | ***0.592*** |
| *Missing obs* | *16* | *5* |  |
| **Mean (SD)** | 576206.62 (1719354.47) | 48925.00 (65442.73) |  |
| **Range** | 286.00 - 7520000.00 | 2650.00 - 95200.00 |  |
| ***Treatment with protease inhibitor*** | | | ***0.687*** |
| *Missing obs* | *1* | *0* |  |
| **No** | 17 (41.5%) | 2 (28.6%) |  |
| **Yes** | 24 (58.5%) | 5 (71.4%) |  |
| ***Absolute CD4 count at diagnosis (cells/mm^3^)*** | | | ***0.756*** |
| *Missing obs* | *11* | *3* |  |
| **Mean (SD)** | 407.39 (306.26) | 372.75 (415.05) |  |
| **Range** | 0.00 - 1016.00 | 4.00 - 800.00 |  |
| ***Interval from diagnosis to ART start (days)*** | | | ***0.815*** |
| *Missing obs* | *5* | *2* |  |
| **Mean (SD)** | 480.11 (1081.91) | 504.60 (579.72) |  |
| **Range** | 0.00 - 5766.00 | 0.00 - 1358.00 |  |
| ***Duration of ART (months)*** | | | ***0.831*** |
| *Missing obs* | *5* | *2* |  |
| **Mean (SD)** | 208.43 (88.07) | 221.60 (67.13) |  |
| **Range** | 32.00 - 382.00 | 124.00 - 305.00 |  |
| ***CD4 nadir (cells/mm^3^)*** | | | ***0.898*** |
| **Mean (SD)** | 226.24 (191.89) | 246.57 (250.75) |  |
| **Range** | 0.00 - 749.00 | 4.00 - 645.00 |  |
| ***Ethnicity*** | | | ***1.000*** |
| *Missing obs* | *3* | *0* |  |
| **Hispanic** | 12 (30.8%) | 2 (28.6%) |  |
| **Not Hispanic** | 27 (69.2%) | 5 (71.4%) |  |

**Extended Data Table 1: Associations between recall cytotoxicity, demographic, and clinical parameters.** Comparison of 16 demographic and clinical parameters between participants with low (AUC <100) and high (AUC >100) HIV-specific CD8^+^ T cell cytolytic capacities as measured in Fig. 2. *p* values were calculated using Fisher’s exact tests for categorical values or Wilcoxon rank-sum tests for continuous variables.

| **H047 Samples** | | |  | | **Days of ATI** |  | **HIV-1 Clinical Viral Load** | | | | **PBMCs** |
| --- | --- | --- | --- | --- | --- | --- | --- | --- | --- | --- | --- |
| TI0 | | |  | | 0 |  | <20; TND | | | | **☑** |
| TI1 | | |  | | 7 |  | <20; TND | | | | ☐ |
| TI2 | | |  | | 14 |  | 34 | | | | **☑** |
| TI3 | | |  | | 18 |  | 250 | | | | ☐ |
| TI4 | | |  | | 25 |  | 33900 | | | | **☑** |
| TI5 | | |  | | 32 |  | 143000 | | | | ☐ |
| TI6 | | |  | | 38 |  | 36500 | | | | **☑** |
| AR2 | | |  | | 50 |  | 34 (on ART) | | | | **☑** |
| AR4 | | |  | | 66 |  | <20; TD (on ART) | | | | **☑** |

**Extended Data Table 2. Viremia as a function of treatment interruption duration.**

TI, treatment interruption; Re-ART, re-initiation of 3-drug antiretroviral therapy; TND, target not detected; TD, target detected. Checkboxes represent time points at which PBMCs were collected for analyses.

**Supplementary Data 1: HEAL participant-level and response-level data.**

Tables reporting individual participant-level (tab 1) and response-level (tab 2) data summarized throughout the manuscript, including HLA genotypes, numbers of HLA-optimal epitopes screened, response breadth, epitope specificities, magnitudes, proliferation, recall cytotoxicity, reservoir metrics, and clinical history parameters.

**Supplementary Data 2: PBMC CITE-seq and immune repertoire analyses.**

Tables reporting PBMC cell numbers sequenced per time point, before and after filtering (tab 1), longitudinal cell type percentages (tab 2), the absolute number of CD8+ T cells per cluster (tab 3), and CD8^+^ T cell clonotype frequencies (tab 4).

**Supplementary Data 3: Differentially expressed gene set subnets**.

Summary of subnets of overlapping Hallmark, gene ontology, reactome, and KEGG gene sets significantly enriched in clusters 0 and 1 via gene set network analysis (GSNA) of HIV-1 Gag AW11-specific CD8^+^ T cells during ATI in participant H047, as summarized in Fig. 4i.
